# Human immunoglobulin gene allelic variation impacts germline-targeting vaccine priming

**DOI:** 10.1101/2023.03.10.23287126

**Authors:** Allan C. deCamp, Martin M. Corcoran, William J. Fulp, Jordan R. Willis, Christopher A. Cottrell, Daniel L.V. Bader, Oleksandr Kalyuzhniy, David J. Leggat, Kristen W. Cohen, Ollivier Hyrien, Sergey Menis, Greg Finak, Lamar Ballweber-Fleming, Abhinaya Srikanth, Jason R. Plyler, Farhad Rahaman, Angela Lombardo, Vincent Philiponis, Rachael E. Whaley, Aaron Seese, Joshua Brand, Alexis M. Ruppel, Wesley Hoyland, Celia R. Mahoney, Alberto Cagigi, Alison Taylor, David M. Brown, David R. Ambrozak, Troy Sincomb, Tina-Marie Mullen, Janine Maenza, Orpheus Kolokythas, Nadia Khati, Jeffrey Bethony, Mario Roederer, David Diemert, Richard A. Koup, Dagna S. Laufer, Juliana M. McElrath, Adrian B. McDermott, Gunilla B. Karlsson Hedestam, William R. Schief

## Abstract

Vaccine priming immunogens that activate germline precursors for broadly neutralizing antibodies (bnAbs) have promise for development of precision vaccines against major human pathogens. In a clinical trial of the eOD-GT8 60mer germline-targeting immunogen, higher frequencies of vaccine-induced VRC01-class bnAb-precursor B cells were observed in the high dose compared to the low dose group. Through immunoglobulin heavy chain variable (IGHV) genotyping, statistical modeling, quantification of IGHV1-2 allele usage and B cell frequencies in the naive repertoire for each trial participant, and antibody affinity analyses, we found that the difference between dose groups in VRC01-class response frequency was best explained by IGHV1-2 genotype rather than dose and was most likely due to differences in IGHV1-2 B cell frequencies for different genotypes. The results demonstrate the need to define population-level immunoglobulin allelic variations when designing germline-targeting immunogens and evaluating them in clinical trials.

**One-Sentence Summary:** Human genetic variation can modulate the strength of vaccine-induced broadly neutralizing antibody precursor B cell responses.

## INTRODUCTION

Prevention of infection in licensed vaccines often correlates with the induction of protective antibodies that have sufficient breadth to cover heterogeneity among circulating strains of the target pathogen (*1-3*). Results from the Antibody Mediated Prevention (AMP) trials provide evidence that infusion with a broadly neutralizing antibody can protect against HIV infection and support induction of bnAbs as an HIV immunization strategy. Germline-targeting vaccine designs are one of several approaches for generating bnAb responses to the highly variable HIV spike protein (*4*). The germline-targeting approach is based on the hypothesis that rare bnAb-precursor naive B cells with well-defined genetic signatures can be activated by a priming immunogen and subsequently matured to bnAb development by heterologous boosting. In this approach, the precursor frequency and binding affinity are critically important and depend on human genetic characteristics.

The IAVI G001 phase 1 clinical trial of eOD-GT8 60mer adjuvanted with AS01_B_ demonstrated for the first time that germline-targeting immunogens can induce bnAb-precursor responses in humans (*4*). The vaccine was administered at weeks 0 and 8. VRC01-class immunoglobulin G (IgG) B cells were found in 35 of 36 vaccine recipients, with median frequencies among memory B cells (MBCs) reaching as high as 0.09% in the low dose group and 0.13% in the high dose group from peripheral blood mononuclear cell (PBMC) samples collected at weeks 4, 8, 10 and 16 (*4*). All post-vaccination MBC samples with VRC01-class B cells had higher VRC01-class frequency than at baseline for the same participant, hence all were regarded as vaccine induced. VRC01-class B cells were detected in two of twelve placebo participants but these were detected both pre- and post-vaccination and hence were regarded as pre-existing VRC01-class memory. A dose effect was observed, with higher VRC01-class median frequencies measured in the high dose group compared to the low dose group at all MBC timepoints, including statistically significant differences at weeks 4 and 16 (*4*). Observation of a dose effect has potential implications for dose selection in future clinical studies and for interpreting mechanisms of germline-targeting priming immunogens. However, IAVI G001 was a placebo-controlled, double-blind, dose-escalation study investigating safety and tolerability, and as such, the two dose arms were sequentially enrolled. Therefore, dose comparisons were susceptible to confounding by unequal distributions of participant characteristics between dose groups that may impact the frequency of targeted bnAb-precursor B cells.

VRC01-class B cells are defined by utilization of an IGHV1-2 *02 or *04 heavy chain and a 5-amino acid light chain CDR3 (*5-11*). In separate work, the IGHV1-2 genotype was determined with nucleotide-level precision for all trial participants in IAVI G001, by generating immunoglobulin M (IgM) libraries from each individual and applying the germline allele inference tool IgDiscover (*4, 12*). In that work, use of the genotype information was limited to showing that the one participant who did not produce VRC01-class responses was the only participant lacking one of the necessary IGHV1-2 alleles (*02 or *04). Here, we looked more deeply at the genotype data, and we found an imbalance in the IGHV1-2 genotype distribution between the low and high dose groups, which suggested that the observed dose effect in VRC01-class responses might have been due at least in part to genotype differences rather than dose. We investigated whether and how the dose level and IGHV1-2 genotype affected the frequency of vaccine-induced VRC01-class IgG B cells, using statistical models tested by independent quantitative analyses of experimentally measured mRNA expression levels and B cell frequencies. We also assessed whether IGHV1-2 alleles were associated with different VRC01-class precursor affinities for the vaccine, to determine if allele effects on affinity could explain the different response rates for the two genotype-unbalanced groups.

## RESULTS

### IGHV1-2 allele frequencies vary by dose group

As reported elsewhere (*4*), through personalized genotyping a total of 9 different IGHV1-2 genotypes were found among the 48 IAVI G001 trial participants (Figs. 1A-B). These consisted of combinations of the known alleles *02, *04, *05 and *06, in addition to a novel allelic variant, IGHV1-2*02_S4953, that is distinct at the nucleotide level but encodes the same amino acid sequence as the *02 allele. For the subsequent analyses in the current study, we therefore classified *02_S4953 as *02. A structural view and amino acid sequence alignment of the various IGVH1-2 alleles is shown in Fig S1. The frequency of each allele in the 48 trial participants showed that IGHV1-2*04 was most common, followed by IGHV1-2*02 (Fig. 1C). The one participant that did not produce a detectable VRC01-class response was found to be IGHV1-2 genotype *05/*06, i.e. lacking one of the required *02 or *04 VRC01-class alleles, at least one of which were present in the genotype of all other trial participants (Fig. 1B and table S1; (*4*)).

**Fig. 1.**
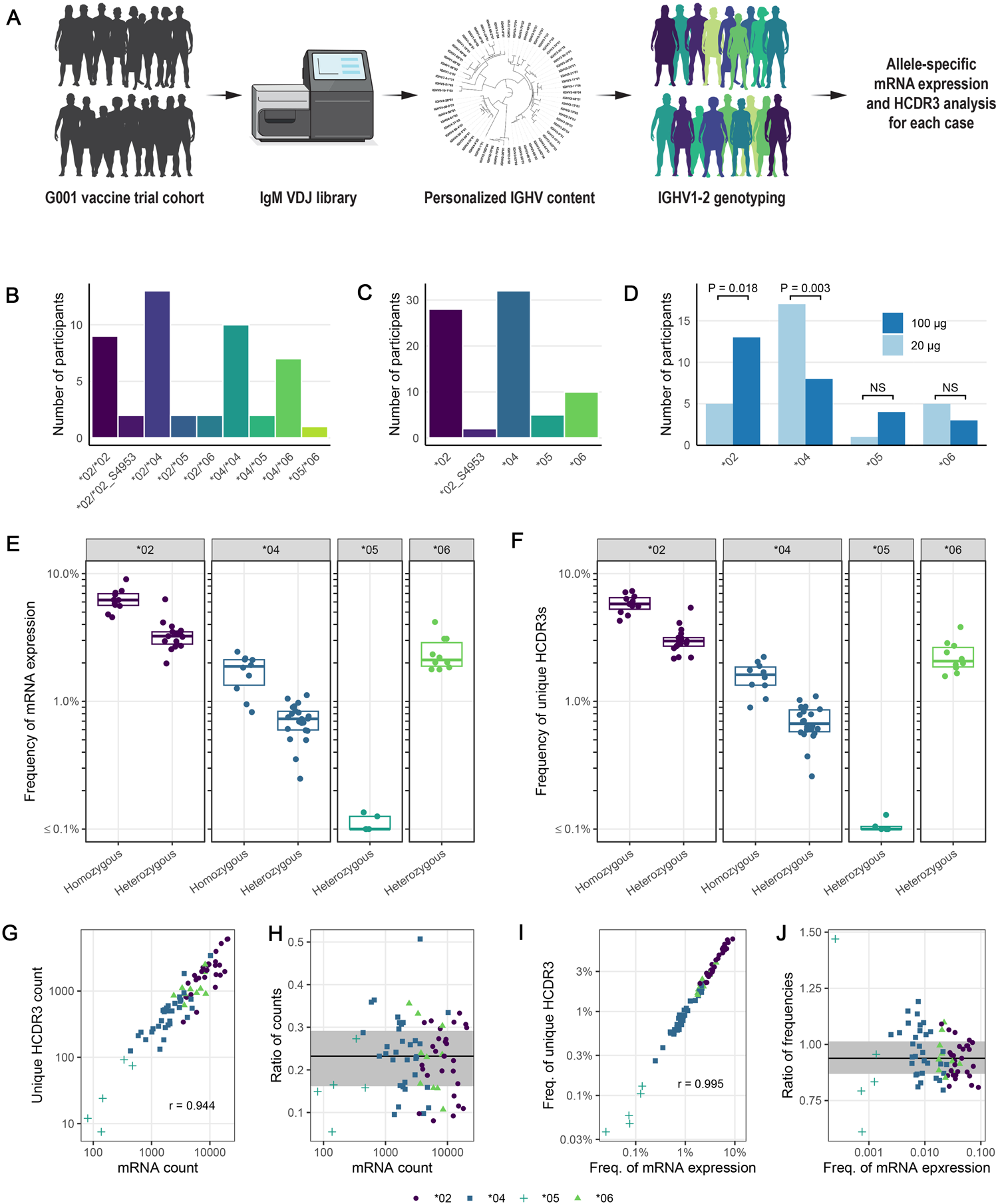
IGHV1-2 genotype, allele, and pre-vaccination IgM repertoire distributions for IAVI G001 trial participants. **(A)** The IGHV1-2 allele content in each study participant was determined by sequencing bulk IgM libraries and inferring the IGHV allele content in each case with IgDiscover. Quantitative analyses of mRNA expression levels and HCDR3 frequencies followed. **(B)** Number of each genotype and **(C)** number of each allele, for all trial participants (n=48). **(D)** Number of vaccine recipients per group, out of 18, with each allele (*02, *04, *05, and *06), with the novel *02 variant *02_S4953 classified as *02. P-values are based on a Fisher’s exact test with values greater than 0.05 marked as not significant (NS). For all 48 participants, pre-vaccination **(E)** mRNA expression frequencies and **(F)** unique HCDR3 frequencies for each IGHV1-2 allele are shown as points color-coded by allele and grouped by homozygous and heterozygous genotype. Each point represents a trial participant, with heterozygous participants represented by two points. Thick lines are median values and boxes are the 25% and 75% quantiles. **(G)** Correlation between mRNA count and unique HCDR3 count; **(H)** Ratio of unique HCDR3 count to mRNA count versus mRNA count; **(I)** Correlation between frequency of mRNA expression and frequency of unique HCDR3s; and **(J)** Ratio of the unique HCDR3 frequency to the mRNA frequency versus the mRNA frequency, are shown for pre-vaccination repertoires. Points are shape- and color-coded by IGHV1-2 allele as shown in the legend of panels G-J. Pearson correlation coefficients (r) for counts and frequencies are shown in panels G and I, respectively. In panels H and J, the solid line is the median ratio, and the shaded region shows the inter-quartile range.

We observed that the distributions of *02 and *04 alleles between dose groups were substantially uneven (Fig. 1D). In the high dose group, 72% of participants (13 of 18) had an *02 allele, compared to 28% (5 of 18) in the low dose group (P-value=0.02). Conversely, in the low dose group, 94% of participants (17 of 18) had an *04 allele, compared to 44% (8 of 18) in the high dose group (P-value=0.003). The *05 and *06 alleles were less prevalent and had similar frequencies between dose groups (Fig. 1D). The imbalance in *02 and *04 between dose groups potentially impacted the relative strengths of the VRC01-class responses, because a previous study had shown that the frequency of eOD-GT8-specific naive precursors in the germline repertoire was higher for individuals encoding the *02 allele compared to those encoding the *04 allele (*13*). Therefore, we hypothesized that the dose effect observed in this trial depended on IGHV1-2 allelic differences between the groups.

### IGHV1-2 allele frequencies differ in the germline repertoire

We calculated the mRNA expression frequencies of different IGHV alleles in the naive repertoire for each trial participant by counting the per-allele unique molecular identifiers (UMIs) introduced during the cDNA synthesis of IgM libraries (Fig. 1A and Data S1). The frequencies for each IGHV1-2 allele are shown separately for homozygous and heterozygous genotypes in Fig. 1E. The mean per-allele mRNA expression of *02 was similar in homozygous and heterozygous individuals, at 3.1% (95% CI: 2.7 to 3.6%) and 3.3% (95% CI: 2.9 to 3.8%), respectively (table S2). Usage of *04 was approximately 4-fold lower, at 0.9% (95% CI: 0.7 to 1.1%) in homozygous participants and 0.7% (95% CI: 0.6 to 0.8%) in heterozygous participants (table S2). Comparison of the per-allele frequencies between homozygous and heterozygous participants suggested that allele usage was proportional to zygosity, with homozygotes having approximately twice the usage of *02 or *04 than heterozygotes (Fig. 1E and table S3). Alleles *05 and *06, which do not have the required VRC01-class binding motif, had very low usage, 0.09% (95% CI: 0.03 to 0.14%), and intermediate usage, 2.4% (95%CI: 1.9 to 3.0%), respectively (table S2). We then counted the number of unique heavy chain complementarity determining region 3 (HCDR3) sequences within IGHV1-2 mRNAs, a measure of the number of unique IGHV1-2 B cells in each personalized library (Figs. 1A and 1F and Data S1). We found that, regardless of IGHV1-2 allele, the number of unique IGHV1-2 HCDR3s was proportional to the number of IGHV1-2 mRNA molecules (Figs. 1G-H), and the frequencies in the repertoire were also proportional (Figs. 1I-J). Indeed, ratios of unique HCDR3 counts to mRNA counts (Fig. 1H), a measure of 1/(BCR cell surface density), were similar for *02 and *04 alleles (median 0.23 and 0.24, respectively; P-value for difference=0.53), which suggests that the surface density was not appreciably different between these two alleles. Additionally, ratios between HCDR3 frequency and mRNA frequency for *02 and *04 (Fig. 1J) were also similar (0.93 and 0.96, respectively; P-value for difference=0.19), which also suggested that surface density was similar in the two cases. Thus, different IGHV1-2 alleles had different frequencies of unique precursor B cells, with *02 higher than *04 (Fig. 1F). Furthermore, *02 or *04 homozygotes had approximately twice the frequency of allele-specific unique precursors as heterozygotes (Fig. 1F). Higher bnAb-precursor frequency has been shown to lead to higher bnAb-precursor-derived vaccine responses in mouse models (*14-16*). Therefore, the higher frequency of unique IGHV1-2 precursors in *02 compared to *04 individuals suggested that the stronger VRC01-class responses in the IAVI G001 high dose group could be due at least in part to the greater representation of *02 in that group. Taken together, these results supported including the *02 and *04 allele counts as independent predictors for the frequency of vaccine-induced VRC01-class IgG B cells in our statistical models.

### One model best explains the difference in VRC01-class responses between dose groups

To look for genotype-specific effects in the vaccine response data, we first analyzed differences in post-vaccination VRC01-class B cell frequencies among trial participants that were *02/*02, *02/*04, *04/*04, or *04 heterozygous with either of the non-productive *05 or *06 alleles, and we found no significant differences after adjusting for multiple comparisons (table S4).

Similarly, we compared across dose groups with sufficient data for two genotypes: i) *02/*04 and ii) *04/*05 or *04/*06 (pooled genotype), and we found that neither comparison was significant (table S5). However, the small numbers of participants within each genotype and dose group meant that the analysis had low sensitivity. To increase sensitivity to detect genotype-specific effects, we used statistical modeling to analyze pooled data under the mechanistic assumption that allele effects were additive.

We modeled the VRC01-class response frequency using four candidate models. Each model was defined by one or more frequency parameters:

- Model 1 (Null): A single frequency for all vaccine recipients.
- Model 2 (Dose): Two frequencies, one for the low dose and another for the difference between high and low dose.
- Model 3 (Allele): Two frequencies, one each for the *02 and *04 alleles.
- Model 4 (Full): Three frequencies, one each for the *02 and *04 alleles at low dose, and another for the allele-independent difference between high and low dose.

As described in Leggat *et al*. (*4*), for all trial participants (n=48), the frequencies of VRC01-class BCRs were measured for three sample types at seven sample collection time points: (i) PBMC IgG memory B cells (MBCs) at weeks 4, 8, 10, and 16; (ii) lymph node IgG germinal center (GC) B cells at weeks 3 and 11; and (iii) PBMC IgD-plasmablasts (PBs) at week 9. Here, we modeled count data of VRC01-class BCRs using a quasi-Poisson generalized linear model. The models were fit separately for each of the seven sample collection time points and ranked based on a Quasi-likelihood version of Akaike’s second-order information criterion (QAICc) (*17*).

The QAICc model selection criterion ranked the models consistently at all four MBC time points, with the following best-to-worst order: (i) Allele; (ii) Full; (iii) Dose; and (iv) Null (fig. S2A and table S6). This model ranking indicated that the differential distributions between the dose groups of IGHV1-2 alleles *02 and *04 better explained the VRC01-class B cell frequencies post vaccination than either dose alone or a simple overall average frequency among vaccine recipients estimated by the Null model. Furthermore, the Allele model ranked higher than the Full model at all MBC time points, which indicated there was no detectable dose effect after accounting for the allele effect. At the week 3 GC and week 9 PB time points, the Allele model was also selected as the best model, but the ranking of the remaining models changed (fig. S2A and table S6). For the week 11 GC samples the Null model ranked highest, suggesting that neither dose nor allele adequately explained the observed variation in those samples.

Parameter estimates from the Allele model were consistently higher for the per-allele contribution of *02 to the VRC01-class response than for the contribution of *04 (fig. S2B and table S7). The Full model estimated similar per-allele effects as the Allele model but also estimated effects for high versus low dose that were close to zero and had confidence intervals that included zero in nearly all cases (fig. S2B and table S7). Thus, the data provided no support for a true dose effect, and these results suggested that the count of *02 and *04 alleles best explained the variation in the VRC01-class B cell response to eOD-GT8 60mer.

We then compared experimentally measured VRC01-class frequencies to the Allele-model-estimated mean VRC01-class responses by genotype (Fig. 2 and table S8). To make comparisons in an ordered manner, we ranked IGHV1-2 genotypes based on the frequency of IGHV1-2 mRNA expression in the naive repertoire: first, we grouped genotypes that had a single *05 or *06 allele together since these alleles lack the necessary VRC01-class motif; next, we ranked the genotypes based on our observation that *02 precursors were approximately four-fold more common than *04 precursors. This resulted in our most-to-least favorable ranking of genotypes for induction of a VRC01-class response: (i) *02/*02; (ii) *02/*04; (iii) *02/*05 or *02/*06; (iv) *04/*04; and (v) *04/*05 or *04/*06. The genotype-specific medians of the experimentally measured VRC01-class frequencies generally followed this same ordering (Fig. 2), and the model-estimated mean VRC01-class response also followed this ordering, except for the week 10 MBC time point where the order of *04/*04 and *02/*05 or *02/*06 was reversed (Fig. 2 and table S8). Hence the model captured the dependence of the VRC01-class response frequency on genotype. However, there was substantial heterogeneity in the responses that was not explained by genotype (Fig. 2). This variation was potentially attributable to other factors that can influence the strength of immune responses, including but not limited to sex, age, additional genetic factors, and immune history. Nevertheless, our modeling indicated that the effect of dose was negligible after accounting for IGHV1-2 allele content.

**Fig. 2.**
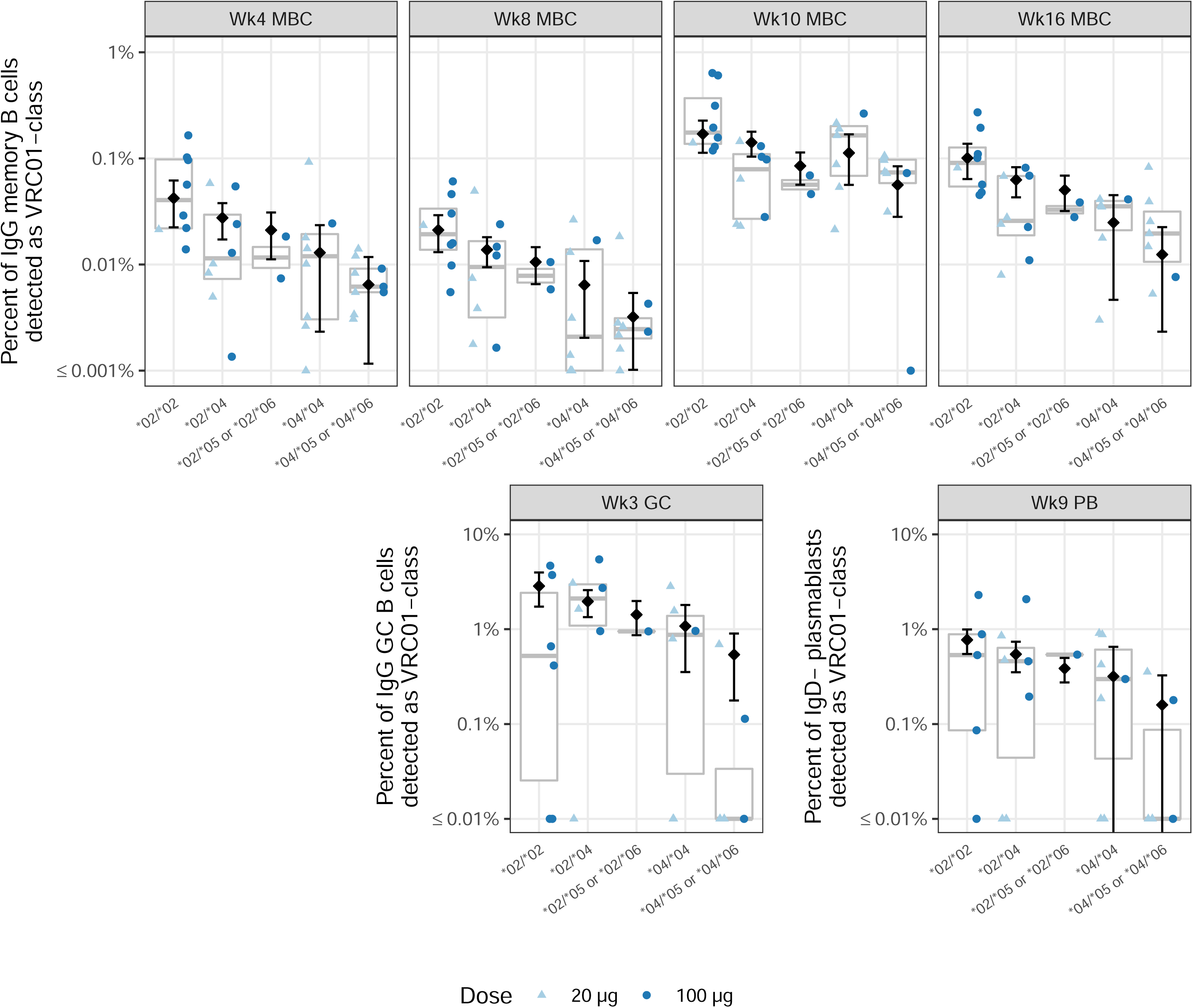
Model estimates and 95% confidence intervals (CIs) from the Allele model for each genotype and time point. Estimates and CIs for the frequency of VRC01-class IgG B cells at each time point by genotype are shown as open diamonds and vertical lines, respectively. Experimentally measured frequencies for each participant are shown as color- and shape-coded points by dose group as indicated by the legend. Genotypes containing the *05 and *06 alleles are grouped together (e.g., *02/*05 or *02/*06), because the estimated mean response from the Allele model depends only on the count of *02 and *04 alleles. Week 11 germinal center (GC) results are not shown since the Null model ranked higher than the Allele model for that sample time point.

### *02 has higher frequencies in both the naive repertoire and the modeled VRC01-class response

From the Allele model, we computed the relative contribution of *02 versus *04 alleles to the post-vaccination VRC01-class B cell frequency at each time point, and we found ratios between 1.5 and 4 (Fig. 3A and table S9). The confidence intervals in all cases included 1.0, consistent with equal contributions from *02 and *04. However, when we computed differences, rather than ratios, in post-vaccination VRC01-class B cell frequencies, we found significantly greater contributions from *02 compared to *04 (table S10; differences significant for all timepoints except week 10). This suggested that the true ratios were greater than 1. In the pre-vaccination IgM (naive) repertoire of vaccine recipients, we computed ratios for *02 versus *04 mRNA usage of 3.9 (95% CI: 3.0 to 5.3) among homozygous individuals and 4.2 (95% CI: 3.3 to 5.1) among heterozygous individuals, both of which were significantly greater than 1.0 (P-values of <0.0001 and 0.0001, respectively) (Fig. 3B and table S11). Ratios of the frequencies of unique HCDR3s using *02 versus *04 in the pre-vaccination repertoire were similar to the mRNA frequency ratios (Fig. 3C and table S11), more directly indicating higher *02 naive B cell frequencies. Overall, mRNA usage and B cell frequency were higher for *02 than for *04 in the personal naive repertoires, and *02 was higher than *04 in the Allele-model-determined contributions to vaccine-induced VRC01-class B cell frequencies.

**Fig. 3.**
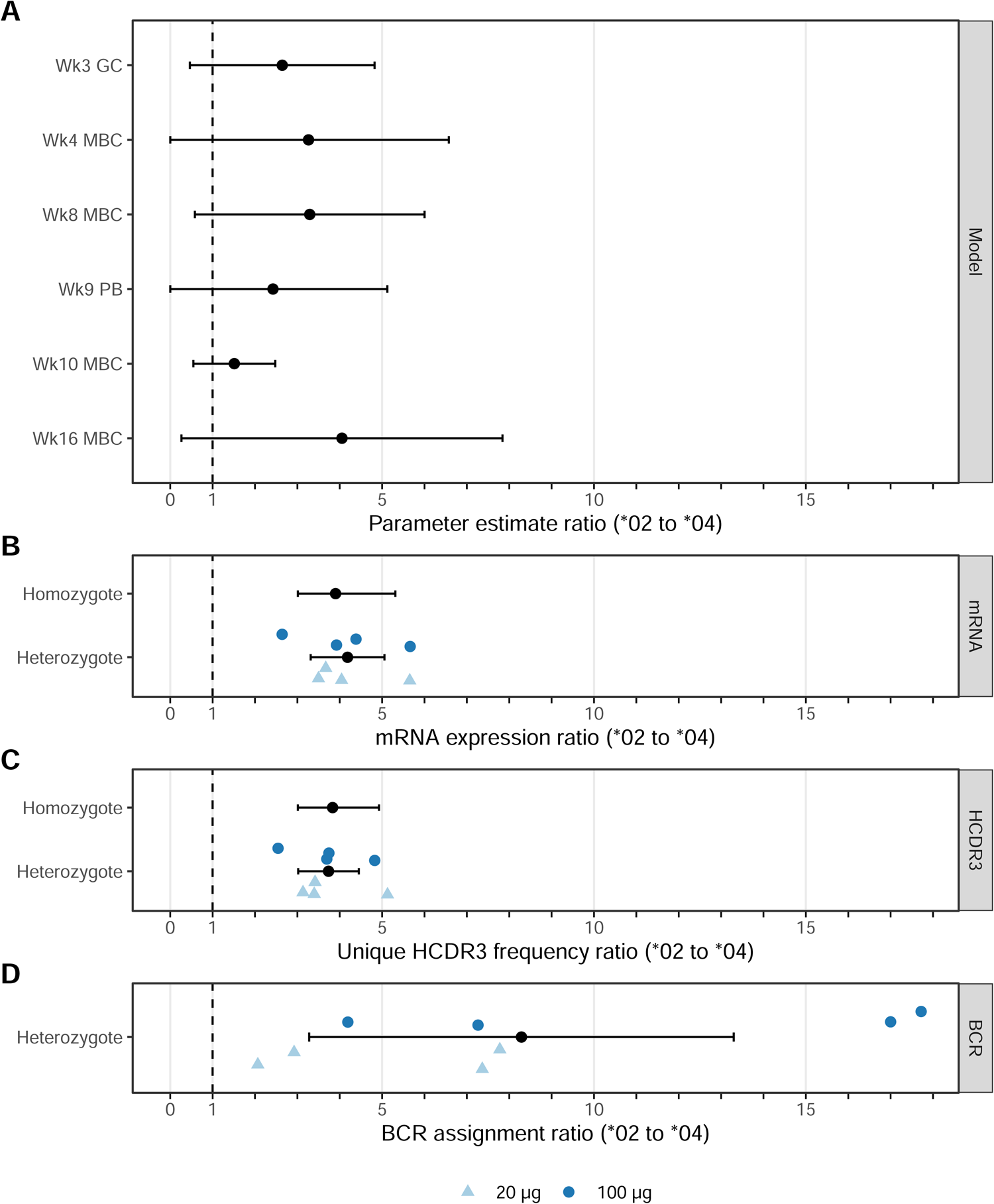
Relative contribution of *02 versus *04 alleles to Allele-model-derived post-vaccination VRC01-class frequency, pre-vaccination IGHV1-2 mRNA expression level, and post-vaccination BCR assignments. **(A)** Allele model estimates for the ratio of the *02 contribution to the *04 contribution to the post-vaccination VRC01-class frequency are shown with 95% confidence intervals (CIs) for germinal center (GC) B cell, memory B cell (MBC), and plasmablast (PB) samples taken at the indicated week (Wk) after first vaccination. **(B)** Experimentally measured pre-vaccination ratios of *02 to *04 mRNA expression levels for homozygous or heterozygous genotypes. For homozygotes, the ratio of means and CI for *02 and *04 individuals is shown. For heterozygotes, ratios for each individual are shape- and color-coded (N = 4 for low dose; N = 4 for high dose), and the overall mean ratio and CI are shown in black. **(C)** Experimentally measured pre-vaccination ratios of *02 to *04 unique HCDR3 frequencies for homozygous or heterozygous genotypes. Homo- and heterozygote data are displayed as in (B). **(D)** Ratio of *02 to *04 usage for germline allele assignments among post-vaccination BCRs recovered from eight vaccine recipients known to be heterozygous for *02 and *04 by pre-vaccination genotyping are shape- and color-coded (N = 4 for low dose; N = 4 for high dose), and the overall mean ratio and CI are shown in black.

### *02 has higher frequencies among allele assignments for post-vaccination BCR sequences

Considering that VRC01-class responses from *02/*04 heterozygous individuals involved direct competition between the two alleles, we assessed frequencies of VRC01-class post-vaccination BCRs with allele assignments of *02 or *04 from *02/*04 heterozygous participants. For each VRC01-class BCR, the IGHV1-2 allele was bioinformatically assigned, accounting for the personal IGHV1-2 genotype information, as described in Willis *et al*. (*5*). From 873 post-vaccination BCR sequences of VRC01-class IgG (MBC or GC) or IgD-(PB) B cells from eight *02/*04 heterozygous vaccine recipients, we computed the per-participant ratio of *02 to *04 as the assigned germline allele (Fig. 3D and table S12). Only four of 873 (0.46%) BCR assignments were ambiguous, hence allele ambiguity did not meaningfully affect our calculations. The overall median ratio of *02 to *04 was 7.3 (range, 2 to 17.7) (table S12), which was significantly greater than 1.0 (P-value = 0.004). The ratio of *02 to *04 BCRs varied widely (0 to infinity) across different post-vaccination time points but was greater than 1.0 in 89% (42/47) of cases (table S13). The observed BCR usage ratios (Fig. 3D and tables S12-S13) were generally higher than the model-derived usage ratios (Fig. 3A and table S9) and the naive repertoire mRNA and HCDR3 frequency ratios (Figs. 3B-C and table S11). We tested for a dose effect in the data in Fig. 3D but found no significant effect (P-value=0.154). IGHV1-2*02 differs from *04 by a single nucleotide (A^*04^/T^*02^, SNP rs112806369) and a single amino acid (Arg_66_^*04^/Trp_66_^*02^, Kabat residue numbering) (*6, 8, 14*). We documented several occurrences of the Arg_66_^*04^⟶Trp_66_^*02^ mutation, which is favorable for VRC01-class maturation, in post-vaccination VRC01-class BCRs from *04 homozygous individuals (*5*). Thus, occurrence of this mutation might have inflated the *02 to *04 ratio in BCRs of heterozygous individuals shown in Fig. 3D. Overall, the *02 to *04 ratios among BCRs from heterozygotes were consistent with the conclusions of the Allele model in that both indicated stronger VRC01-class responses from the *02 allele. Those findings, combined with the fact that the naive repertoire mRNA and HCDR3 ratios demonstrated higher expression and B cell frequency for the *02 allele, suggested a simple potential explanation for the superiority of *02 over *04 for VRC01-class responses, namely that the higher frequency of *02-using B cell precursors translated into higher post-vaccination VRC01-class responses.

### Naive repertoire predicts outcome

To test the hypothesis that stronger VRC01-class responses resulted from higher B cell precursor frequencies, we looked for correlations between the VRC01-class response and the total frequency of IGHV1-2*02 or *04 B cells in the naive repertoire. Pooling across dose groups (Fig 4), we found significant correlations at each time point, excluding week 11, with correlation coefficients ranging from 0.4 to 0.6 (Fig.4, P-values ranging from 0.04 to 0.0003). Analyzing dose groups separately, both dose groups showed positive correlations, but the correlations were stronger and statistically significant only for the high dose group (fig. S3). The difference in correlation strength could reflect a dose effect but could also be explained by the fact that the high dose group, which was over-represented by *02, generally had stronger VRC01-class responses with higher dynamic range, and had higher standard deviation of HCDR3 frequency perhaps due to its wider range of genotypes. Whatever the explanation, the significant positive correlations in the pooled data demonstrated that the strength of the VRC01-class response increased monotonically with the total frequency of IGHV1-2*02 and *04 naive precursors. This finding, together with the observation that the experimentally measured naive precursor frequency was higher for *02 than for *04, indicated that the stronger VRC01-class responses for *02 were most likely due simply to higher naive B cell frequencies for *02. Thus, experimental data provided independent corroboration for our statistical modeling.

**Fig. 4.**
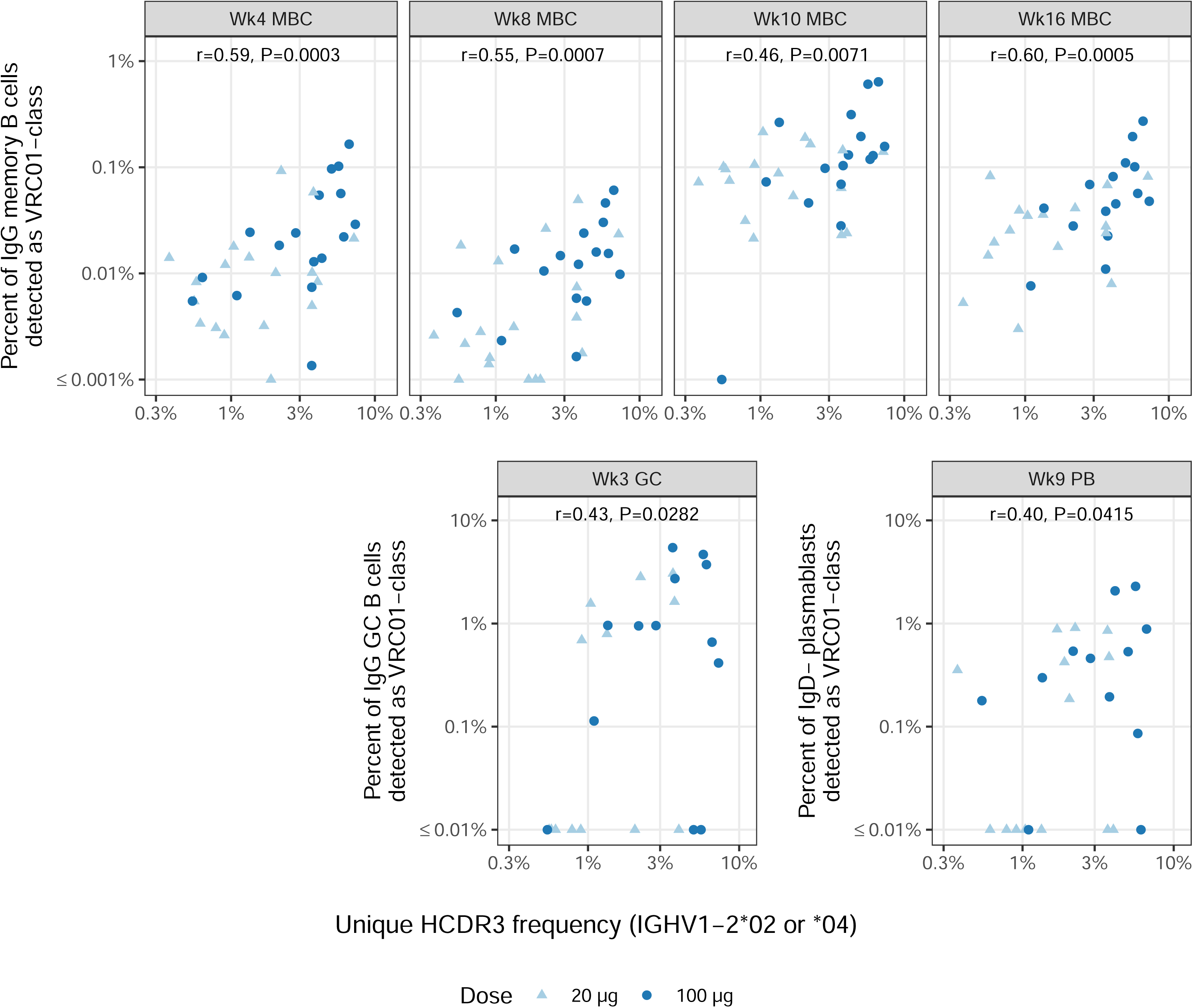
Correlations between pre-vaccination IgM unique HCDR3 frequency (IGHV1-2*02 or *04) and the percent VRC01-class B cell response by visit. Points are shape- and color-coded as shown in the legend. Spearman correlation coefficient (r) and P-values are displayed for each time point. At week 11 in GC the correlation was low (r=0.24, P=0.2287; data not shown).

### Alleles affect precursor affinities

The fundamental hypothesis of the germline-targeting priming strategy is that vaccine antigen affinity and avidity for rare bnAb-precursor B cells strongly impacts whether or not the vaccine can trigger GC and memory responses from those precursors. Hence, while our above analyses focused on precursor frequency, it was also important to consider whether affinity differences between alleles could explain the different response outcomes. To begin, we investigated the effect of the IGHV1-2 *05 and *06 alleles on the affinity of eOD-GT8 for VRC01-class precursors. Alleles *05 and *06 both possess Arg^50^ instead of Trp^50^, one of the critical VRC01-class germline residues that interacts with HIV gp120. We produced W50R variants of VRC01-class precursors that were originally *02 or *04, including inferred germline (iGL) precursors for two bnAbs (VRC01 and N6), iGLs for four post-vaccination BCRs in the IAVI G001 low dose group (*4*), and four human naive precursors isolated by prior B cell sorting studies of HIV-unexposed individuals (*18, 19*). We then assessed eOD-GT8 binding affinity for the original and W50R-variant iGL precursors using surface plasmon resonance (SPR). The original precursors all bound to eOD-GT8, with a median KD of 120 nM, whereas only one of the W50R variants had detectable affinity, and the overall median K_D_ was ≥100μM (Fig. 5A and Data S2). The only W50R binder (K_D_, 4.0 μM) derived from the highest affinity original Ab, VRC01 iGL (K_D_, 49 pM), with an approximately 80,000-fold loss in affinity due to W50R. These results provided an explanation for why eOD-GT8 60mer failed to induce VRC01-class responses in the one *05/*06 participant.

**Fig. 5.**
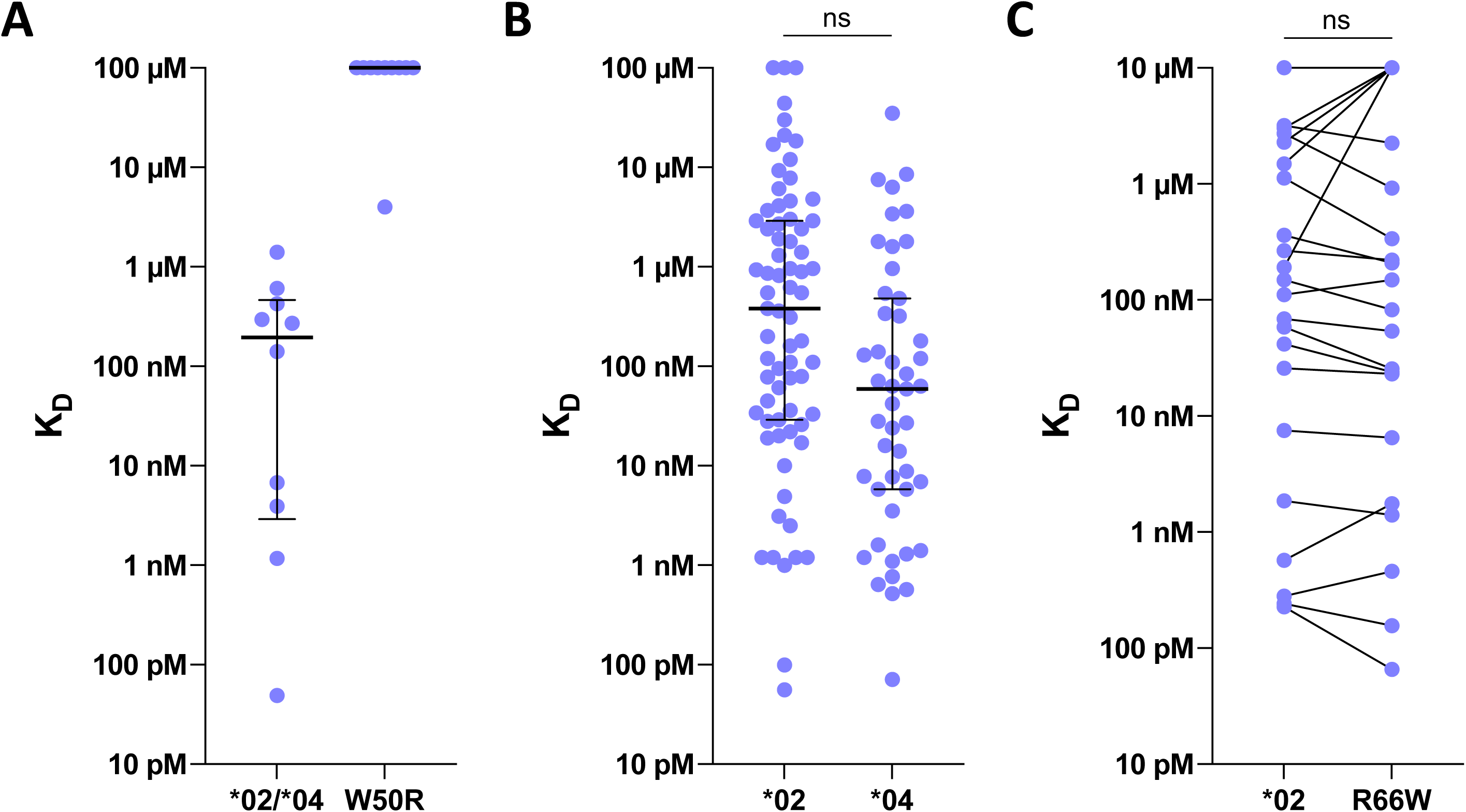
Affinity analyses of VRC01-class IGHV1-2 allele variants. **(A)** Affinities of VRC01-class precursors from two bnAb iGLs, four human naive precursors, and four iGLs from week 3 post-vaccination BCRs from IAVI G001, with original *02 or *04 Abs on the left, and W50R variants of those Abs on the right. **(B)** Affinities of *02 (N=71) and *04 (N=47) iGLs from post-vaccination BCRs from IAVI G001. **(C)** Affinities of *02 VRC01-class iGL antibodies from IAVI G001 and for *04 (R66W) variants of the same antibodies (N=28 each). Lines connect matched Ab variants. In (A) and (B), horizontal lines indicate median and interquartile range. In (A)-(C), all affinities were measured for eOD-GT8 monomer analyte, and all iGLs from IAVI G001 are from the low dose group.

To complement our findings above that VRC01-class *02 precursors had higher frequencies than *04 in IAVI G001, we evaluated whether the two precursor populations had differing affinities for eOD-GT8. We first re-analyzed the Leggat *et al*. data on eOD-GT8 affinities for inferred germline (iGL) variants of post-vaccination VRC01-class BCRs recovered from the low dose group. These iGLs were overwhelmingly (96%) derived from memory BCRs after the first or second vaccination, hence they represented precursors to vaccine responses that survived GC competition and became memory B cells in the blood. Leggat *et al*. reported statistics on all VRC01-class iGLs (median K_D_, 119 nM; N=118), but here we examined the data for *02 and *04 iGL precursors separately. We found that *04 iGLs, with median K_D_ of 59 nM and interquartile range of 5.8 nM to 480 nM (N=47), had approximately six-fold higher affinities than *02 iGLs (median K_D_, 380 nM; interquartile range, 29 nM to 2.9 μM; N=71) (Fig. 5B and Data S2). However, using a repeated measures test to account for the fact that affinities for different antibodies from the same participant are potentially correlated (the N=47 *04 Abs came from 14 participants, and the N=71 *02 Abs came from 5 participants), the six-fold difference was not significant (p-value, 0.38). Furthermore, we tested directly whether *04 precursors had inherently higher affinity than *02 precursors as a consequence of the R66W mutation that distinguishes *02 (Arg^66^) from *04 (Trp^66^). Although this mutation is not located within the region of direct antibody-antigen contacts, the spatial location of this mutation suggested that an indirect (allosteric) effect on affinity was possible (fig. S1). To test for such an effect, we produced 28 different *02 VRC01-class iGL antibodies from the IAVI G001 low dose group, along with *04 (R66W) variants of the same antibodies, and we measured their affinities for eOD-GT8 monomer by SPR (Fig. 5C and Data S2). We found that the original *02 iGLs (median K_D_, 310 nM; interquartile range, 30 nM to 8.3 μM; N=28) had indistinguishable affinities to their *04 variants (median K_D_, 280 nM; interquartile range, 23 nM to 10 μM; N=28; repeated measures test p-value, 0.64). We concluded that neither allele had a precursor affinity advantage over the other. Therefore, affinity differences between alleles could not explain the different response outcomes between the vaccine groups.

## DISCUSSION

The IAVI G001 clinical trial established proof of concept for the germline-targeting strategy of priming bnAb-precursor B cell responses (*4*). In this study we extend those results to show how IG genotype influenced the human response to the vaccine used in the trial. Higher frequency VRC01-class responses were observed in the high dose compared to the low dose group, but here, through quantification of personalized allele usage, HCDR3 frequencies, and allele-specific mAb affinities, and statistical analyses of genotype effects, we have demonstrated that the apparent dose effect is best explained by an imbalance in IGHV1-2 alleles between dose groups. The results of this study, while not precluding that there could be a true dose effect within genotype, show that functionally consequential allelic variation can impact the performance of germline-targeting vaccine priming on at least two levels. First, particular germline alleles can dramatically alter affinity for the priming immunogen and thereby act as an on-off switch for the desired response, as demonstrated by the >4-orders of magnitude loss of eOD-GT8 affinity for precursors with the W50R mutation present in the *05 and *06 alleles, and the corresponding absence of VRC01-class responses in the *05/*06 heterozygous genotype participant in contrast to all other participants. Second, different alleles can have different utilization frequencies within the naive repertoire, and corresponding different B cell frequencies, which can translate directly into different vaccine response frequencies, as demonstrated by the highest responses being consistently found in cases that contained at least one *02 allele compared to those that did not have any *02 allele. Our study supports the idea that germline-targeting approaches in general should consider both the affinity effects and the relative abundances of the targeted alleles in the naive repertoire, both of which are dependent on the genotype. Furthermore, imbalances across groups in the allele distributions of the targeted bnAb-precursor in a trial of a germline-targeting vaccine should be accounted for when making group comparisons. For clinical tests of germline-targeting immunogens, our results encourage a practice of genotyping of trial participants for antibody genes relevant to the antibody-antigen interaction in question. The genotype information should be used either during randomization into groups, to ensure a balanced distribution of relevant alleles in advance, or during analysis of trial results in concert with statistical modeling, mRNA usage quantification, and antibody affinity analysis approaches described here, to ensure that allele-specific effects can be controlled. Overall, our results emphasize the importance of accounting for allele content and the frequencies of these alleles in the naive repertoire when developing and analyzing germline-targeting vaccines.

Our results also provided evidence that precursor frequency affects the ability of germline-targeting priming immunogens to induce bnAb-precursor-derived responses in humans. In mouse models, it has been established that the performance of a germline-targeting immunogen depends on at least three factors: 1) the target precursor B cell frequency, 2) the monovalent affinity of the precursor B cell to the immunogen, and 3) the avidity or multivalency of the immunogen (*5, 14-16, 18, 20*). The eOD-GT8 60mer immunogen was designed to achieve high affinity and avidity (criteria 2 and 3) for diverse VRC01-class precursors, with the assumption that targeting diverse precursors would increase the target precursor B cell frequency (criteria 1) and would be needed to target a sufficiently large pool of precursors in any individual at any one time. Human genetic diversity and antibody recombinational diversity underlie the assumed need to target diverse precursors sharing a minimal required set of bnAb characteristics (*18, 21, 22*). We found that the frequency of IGHV1-2*02 mRNA usage was approximately four-fold higher than for the *04 allele, and correspondingly, we found that the *02 naive B cell frequency was approximately four-fold higher than for the *04 allele, consistent with a previous observation of higher frequencies of eOD-GT8-specific VRC01-class precursors in *02 compared to *04 individuals (*13*). We also showed that neither allele had an affinity advantage. Thus, increased precursor frequencies in *02 individuals provide a simple potential mechanistic explanation for the elevated post-vaccination VRC01-class frequencies we observed here in *02 compared to *04 participants. Taken together, these results provide evidence that bnAb precursor frequency (criterion 1) is also important in humans, and that individual-to-individual variation in VRC01-class precursor frequency was controlled by IGHV1-2 genotype variation in this trial.

Our findings underscored the utility of statistical modeling for identifying mechanistic effects in clinical data. The IAVI G001 clinical study was not originally designed to study genotype-specific effects, and consequently, comparisons between VRC01-class responses for different genotypes at any one time point were not sufficiently powered to detect differences. However, in our models we used the simplifying assumption that each allele contributes to the VRC01-class response independently, which allowed us to estimate *02 and *04 allelic effects using data from nearly all vaccine recipients. At most time points, we found that *02 and *04 allele content alone best predicted the VRC01-class response; that is, allelic content predicted the vaccine-induced response better than dose, and after adjusting for allelic content, dose did not sufficiently explain the remaining variation in the vaccine-induced response to warrant inclusion in the best model.

We therefore established that trial participant IGHV1-2 allele content confounded the relationship between dose and the vaccine response. We believe this is the only known example of an established confounder of the dose effect found in a clinical dose-escalation vaccine trial. A Pubmed search using the search terms “confound”, “vaccine”, and “dose escalation” resulted in two references (*23, 24*), neither of which discussed confounding of dose with a baseline participant characteristic.

These results add to a growing body of work investigating the importance of germline variation in immunoglobulin genes and the role of this variation in antibody response to various targets. Early studies of allele effects on antibodies against *Haemophilus influenzae* type b (Hib) dependent on either IGHV3-23 or IGK2-29 revealed geographic and ethnic population-dependent variations in polymorphisms that can affect antibody affinity or expression levels, and also showed that copy number variations that could affect expression levels (*25-28*). The structural interaction of anti-influenza IGHV1-69 bnAbs with the Hemagglutinin stem depends in part on Phe^54^ (*29-32*), a residue encoded in most but not all VH1-69 alleles, with other alleles having Leu^54^ (*28, 33*). Polymorphism at IGHV1-69 position 54 has been shown to impact responses to the influenza hemagglutinin stem epitope during infection or vaccination, with higher germline affinities usually but not always associated with Leu^54^ (*29, 33, 34*), and stronger memory B cell responses and serum antibody binding responses occurring for Phe^54^ homozygotes than Leu^54^ homozygotes, potentially due to preferred IGHV1-69 germline gene usage for Phe^54^ alleles (*33-35*). Different classes of HIV bnAbs derive from different antibody genes (*36*), and the allelic dependencies in most cases remain to be elucidated. The considerations and methods employed in our study should assist further exploration of genotype effects on antibody immunity.

Our results suggest that the lower dose could be used in future studies of eOD-GT8 60mer/AS01_B_, based on: (i) our analyses showing lack of a true dose effect; (ii) the separate demonstration of favorable affinity maturation of VRC01-class compared to non-VRC01-class BCRs studied for the low dose group (*4*), which in light of our present analysis shows that the low dose responses were highly productive even in cases with a genotype disadvantage; and (iii) the general consideration that the lowest safe and effective dose should be used. Furthermore, given that germline-targeting priming immunogens are designed with the intention of providing a competitive advantage to bnAb-precursor B cells, intuitively one would expect that reducing the dose might improve performance by retaining bnAb-precursor activation while reducing non-specific activation of competitors.

Despite the fact that VRC01-class responses were weaker in *04 compared to *02 individuals, we note that VRC01-class response rates and frequencies were nevertheless high among *04 homozygous individuals (e.g., 100% positivity and median frequency of 0.16% among MBCs at week 10; N=7) and even among *04/*05 or *04/*06 heterozygous individuals (e.g. 88% positivity and median frequency of 0.07% at wk 10; N=8) (*4*). Thus, the vaccine was able to induce strong VRC01-class IgG B cell responses regardless of IGHV-1 genotype, for individuals with at least one required allele (*02 or *04). Given that approximately 98.4% of humans are at least heterozygous for *02 or *04 (*13*), our analyses of the IAVI G001 trial provide further encouragement for attempts to develop a vaccine to induce VRC01-class bnAbs. The implications of this study are not limited to IGHV1-2, however. Germline-targeting vaccines targeting other bnAbs for HIV or other pathogens will depend on other human antibody gene variants. This study illustrates for the first time how immunoglobulin genotype can modulate human vaccine responses and how personalized immunoglobulin genotyping can be employed to control for and potentially predict such effects.

## MATERIALS and METHODS

### Study design

IAVI G001, with ClinicalTrials.gov registry number NCT03547245, was a phase 1, randomized, double-blind, placebo-controlled dosage escalation study to evaluate the safety and immunogenicity of eOD-GT8 60mer vaccine adjuvanted with AS01B in HIV-uninfected, healthy adult volunteers. Additional details are in Leggat *et al*. (*4*). Here, we employed immunoglobulin heavy chain variable (IGHV) genotyping, statistical modeling of VRC01-class vaccine responses, quantification of IGHV1-2 allele usage and B cell frequencies in the naive repertoire for each trial participant, and SPR analyses of antibody affinities to investigate a potential genetic explanation for the observed stronger VRC01-class responses in the high dose group of the trial. We assessed the dependence of post-vaccination VRC01-class B cell frequencies on IGVH1-2 genotype alone, dose alone, or genotype and dose, and on pre-vaccination naive repertoire IGVH1-2 allele mRNA expression frequencies and unique precursor B cell frequencies. Additionally, we assessed in vivo competition between IGHV1-2 alleles by evaluating post-vaccination BCR IGHV1-2 gene assignments in VRC01-class responses.

### B cell sorting and sequencing and inferred germline analysis

The primary immunogenicity readout in the trial, frequency of VRC01-class IgG B cells, was measured by fluorescence-activated cell sorting (FACS) and single B cell sorting, B cell receptor sequencing, and bioinformatic analysis, as described in Leggat *et al*. (*4*).

### NGS library preparation

The procedures described in Leggat *et al*. (*4*) and (*37*) were utilized in NGS library preparation. In brief, following cDNA synthesis with an IgM-specific primer that contained a Unique Molecular Identifier (UMI) and a universal amplification sequence, two independent IgM libraries were prepared for each trial participant (N=48). The first utilized a 5’ multiplex set of primers that target all functional IGHV leader regions (leader library) and the second a 5’ multiplex set that targeted the 5’ UTR of the same genes (5’UTR library). In both cases the universal reverse primer was used as the reverse primer.

### Genotype analysis and relative IGHV mRNA quantification

The amplified libraries were individually indexed and sequenced using the Illumina MiSeq platform version 3 (2 × 300 cycle) kit to enable full VDJ coverage. IGHV germline inferral using IgDiscover (version 0.12.4.dev266+ge9e3119) was performed to identify the genotype of each case, as described in (*37*) using the default parameters and the IMGT human IGH database, downloaded in April 2021, as a starting reference database. In addition to producing an individualized genotype for each case, the output of the program provided a count of the number of unique UMIs associated with sequences with zero differences from the inferred individualized germline set. This enabled the UMI count of unmutated sequences to be utilized as a means of calculating overall frequency of unmutated germline alleles in the mRNA molecules used to produce the library. The number of unique UMIs associated per allele of the individualized genotype was used to calculate the overall frequency of different IGHV alleles in the full naïve repertoire in the 5’UTR libraries and the leader libraries of each case in this study and to calculate the relative frequency of different alleles of the IGHV1-2 gene.

### IGHV1-2 HCDR3 frequency quantification

The IgDiscover program determined the number of unique HCDR3s present in the full germline set of UMI containing unmutated IGHV allelic sequences. This enabled the calculation of the frequency of unique HCDR3s associated with each IGHV germline allele present in the 5’UTR IgM libraries and the IgM leader libraries of each case.

### IGHV1-2 allele usage analysis

For each participant allele, the relative usage for that allele was the mean frequency of the two primer sets. For two participants with *02/*02_S4953 genotype, we reported allele usage for *02 as the sum of the mean frequencies from the two primer sets for the *02 and *02_S4953 alleles. As noted in Leggat *et al*. (*4*), the *02_S4953 allele is a novel variant of *02 with a non-coding polymorphism and a similar relative frequency as the *02 allele.

### Statistical modeling analysis

At each of the seven sample collection time points, the frequency of VRC01-class IgG B cells was estimated by taking the product of the frequency of epitope-specific IgG B cells measured using FACS and the frequency of VRC01-class IgG B cells among successfully sequenced epitope-specific B cells measured by B cell sorting, sequencing and bioinformatic analysis, as described in Leggat *et al*. (*4*). We estimated the total number of VRC01-class IgG B cells (V) in each sample by taking the frequency estimate and multiplying by the number of VRC01-class IgG B cells rounded to the nearest integer. We modeled the count data V along with the total number of IgG B cells (N) using an over-dispersed Poisson (or quasi-Poisson) distribution, a relaxed Poisson distribution that allows for the variance to be greater than the mean (*38*).

Estimation was performed using the method of quasi-likelihood (*39*). We defined four models (Null, Dose, Allele, and Full) for each time point given an expectation formula describing the relationship between V and N and the following participant level covariates:

1. 𝕀_*Dose*=100*μg*_ an indicator of being in the high dose group.
2. *n*02 the IGHV1-2*02 zygosity (0, 1 or 2).
3. *n*04 the IGHV1-2*04 zygosity (0, 1, or 2). where the four expectation formulas (identified by the name in parentheses) are:
4. (Null) *E*(*V*|*N*) = β_lntercept_N
5. (Dose) *E*(*V*|*Dose, N*) = β_Dose=20μg_ *N +* β_*Dose delta* (100*μg*-20*μg*)_ · 𝕀_*Dose*=100*μg*_ · N
6. (Allele) *E*(*V*|*n*02, *n*04, *N*) = β *_02_n_02_N+ β *_04_*n*_04_*N*
7. 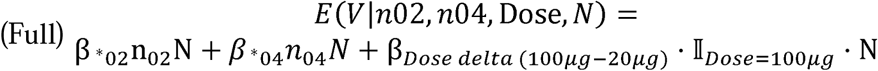

Each model was defined by one or more betas that defined a population average frequency of VRC01-class response among a group of vaccine recipients. The following table describes the model(s) that beta appears in and the interpretation. Figures that show these beta estimates use are annotated using the subscript.

| Model(s) | Beta | Interpretation |
| --- | --- | --- |
| Null | $\beta_{Intercept}$ | Overall vaccine-induced mean response pooling over dose groups |
| Dose | $\beta_{Dose=20\mu g}$ | Mean response in the low dose group |
| Dose, Full | $\beta_{Dose\ delta\ (100\mu g-20\mu g)}$ | Difference in mean response between dose groups |
| Allele, Full | $\beta_{*02}$ | Mean IGHV1-2*02 per-allele response |
| Allele, Full | $\beta_{*04}$ | Mean IGHV1-2*04 per-allele response |

At each time point these models were ranked using the Quasi-likelihood version of Akaike’s second-order information criterion (QAICc) (*18*).

### Bioinformatic BCR sequence analysis

BCR IGHV1-2 allele assignments were determined using Sequencing Analysis and Data library for Immunoinformatics Exploration (SADIE) as described in Willis *et al*. (*5*). For purposes of defining the ratio of *02 to *04 assignment, ambiguous BCR assignments were given 0.5 counts to each allele.

### Antibody production for SPR

Genes encoding the antibody Fv regions were synthesized by GenScript and cloned into antibody expression vectors pCW-CHIg-hG1, pCW-CLIg-hL2, and pCW-CLIg-hk. Monoclonal antibody production was conducted in house using transient transfection of HEK-293F cells (ThermoFisher) and purification using rProtein A Sepharose Fast Flow resin (Cytiva).

### Antigen production for SPR

His-tagged and avi-tagged monomeric eOD-GT8 was produced by transient transfection of HEK-293F cells (ThermoFisher) and purified by immobilized metal ion affinity chromatography (IMAC) using HisTrap excel columns (Cytiva) followed be size-exclusion chromatography (SEC) using Superdex 75 10/300 GL (Cytiva).

### SPR

The data in Figure 5B was measured in Leggat et al. (*4*), while the data in Figures 5A and 5C were measured in this work. In all cases, we measured the kinetics and affinities of antibody-antigen interactions on a Carterra LSA instrument using HC30M or CMDP sensor chips (Carterra) and 1x HBS-EP+ pH 7.4 running buffer (20x stock from Teknova, Cat. No H8022) supplemented with BSA at 1 mg/ml. We followed Carterra software instructions to prepare chip surfaces for ligand capture. In a typical experiment, approximately 2500 to 2700 RU of capture antibody (SouthernBiotech catalog # 2047-01) in 10 mM Sodium Acetate pH 4.5 was amine coupled. The critical detail here was the concentration range of the amine coupling reagents and capture antibody. We used N-Hydroxysuccinimide (NHS) and 1-Ethyl-3-(3-dimethylaminopropyl) carbodiimidehydrochloride (EDC) from Amine Coupling Kit (GE order code BR-1000-50). As per kit instruction 22-0510-62 AG, the NHS and EDC should be reconstituted in 10 ml of water each to give 11.5 mg/ml and 75 mg/ml respectively. However, the highest coupling levels of capture antibody were achieved by using 10 times diluted NHS and EDC during surface preparation runs. Thus, in our runs the concentrations of NHS and EDC were 1.15 mg/ml and 7.5 mg/ml. The concentrated stocks of NHS and EDC could be stored frozen in -20C for up to 2 months without noticeable loss of activity. The SouthernBiotech capture antibody was buffer exchanged into 10 mM Sodium Acetate pH 4.5 using Zeba spin desalting columns 7K MWCO 0.5ml (catalog # 89883 from Thermo) and was used at concentration 0.25 mg/ml with 20 minutes contact time. Phosphoric Acid 1.7% was our regeneration solution with 60 seconds contact time and injected three times per each cycle.

Solution concentration of ligands was around 5 ug/ml and contact time was 3 min. Raw sensograms were analyzed using Kinetics software (Carterra), interspot and blank double referencing, Langmuir model. Analyte concentrations were quantified on NanoDrop 2000c Spectrophotometer using absorption signal at 280 nm. A typical SPR run tested 6 different analyte concentrations using a dilution factor of 4. Maximum analyte concentration for eOD-GT8 was 87 μM for Fig. 5A and 10 μM for Fig. 5C. For the data from Leggat et al. (*4*) in Fig 5B, maximum analyte concentration was generally 10 μM, except for weak binders which were generally re-run at higher maximum analyte concentrations of 50 or 118 μM.

### Statistical analysis

A Fisher’s exact test was used to compare the distribution of IGHV1-2 alleles between dose groups (Fig. 1D). For alleles *02 and *04, the distribution of the ratios of unique HCDR3 counts to mRNA counts (Fig. 1H) and the ratios of HCDR3 frequency to mRNA frequency (Fig. 1J) were compared using a Wilcoxon rank sum exact test. Estimates and 95% confidence intervals (CIs) for both genotype-specific effects and the difference between the *02 and *04 per-allele effect were computed using the *lincom* function in R based on the associated linear combinations of the *02 and *04 coefficients (Fig. 2 and table S8). Ratio estimates and 95% CI for the relative usage of *02 and *04 from the Allele model were computed using the delta method (*40*) truncating the lower bound at zero (Fig. 3A and table S9). The 95% CI for the mean ratios of *02 to *04 mRNA expression, unique HCDR3 frequency, and BCR assignments among heterozygous vaccine recipients were computed using a t-distribution (Fig. 3B-D and table S11). The associated P-values for these ratios were computed using a t-test of the null hypothesis that the ratio is equal to one (Fig. 3D and table S11). The 95% CI and P-value for the ratio of means between homozygous *02 and homozygous *04 vaccine recipients homozygous were computed based on 10,000 bootstrap samples (Fig. 3B-C). The 95% CI and P-value for per-allele differences in mean mRNA expression between homozygous and heterozygous participants were computed using a t-test under the null hypothesis that the difference was zero (table S3).

Comparisons of post-vaccination VRC01-class B cell frequencies between pairs of genotypes were computed using a Wilcoxon rank sum test (table S4). Regarding the analysis of affinity differences between populations of iGL antibodies in Figures 5B and 5C, we observed an association of iGL affinity with participant ID, hence we concluded that applying tests that assume independence of all affinities would be inappropriate. To analyze the difference in mean affinity between *02 and *04 iGL Abs from Leggat *et al*. (Fig. 5B), we used a linear mixed effects model (LME) with fixed effects for the affinity for each allele, and a random intercept to account for within-participant correlations. To analyze the affinity difference between original *02 iGLS and their matched R66W (*05) variants (Fig. 5C), we used an LME with a fixed effect for the difference and a random intercept to account for within-participant correlations. LMEs were fit and p-values were evaluated using Satterthwaite’s degrees of freedom method using the lmerTest package in R (*41*). The R language and tidyverse R packages were used for graphical and statistical analysis (*42, 43*). Graphpad Prism was also used for Figure 5.

## Data Availability

All data are available in the main text or supplementary materials

## List of Supplementary Materials

Fig. S1 to S3

Tables S1 to S13

Data S1 to S2

## Acknowledgments

We thank Pervin Anklesaria, Nina Russell, and Emilio Emini for discussions and trial planning; Jeong Hyun Lee for comments on the manuscript; Olayinka Fagbayi and Amelia Mosley of IAVI for project management at the IAVI NAC at Scripps.

## Funding

This work was supported by the Bill and Melinda Gates Foundation Collaboration for AIDS Vaccine Discovery (CCVIMC INV-007371 to R.A.K., A.B.M., and M.J.M.; VISC INV-008017 and INV-032929 to A.C.D.; VxPDC INV-008352 and INV-007375 to IAVI; and NAC INV-007522 and INV-008813 to W.R.S.), IAVI (including IAVI 167627819 to M.J.M. and other support to W.R.S.), the IAVI Neutralizing Antibody Center (NAC) to W.R.S., National Institute of Allergy and Infectious Diseases (NIAID) P01 AI094419 (HIVRAD Optimizing HIV immunogen-BCR interactions for vaccine development”) (to W.R.S.), UM1 Al100663 (Scripps Center for HIV/AIDS Vaccine Immunology and Immunogen Discovery) and UM1 AI144462 (Scripps Consortium for HIV/AIDS Vaccine Development) (to W.R.S. and M.J.M.); and UM1AI069481 (Seattle-Lausanne CTU), U19AI128914 (HIPC), and UM1AI068618 (HVTN LC) to M.J.M.; and by the Ragon Institute of MGH, MIT, and Harvard (to W.R.S.).

## Author contributions

W.J.F., M.M.C, J.R.W., A.C.D., G.B.K.H. and W.R.S. designed the study. C.A.C., D.L.V.B., and W.R.S. designed SPR studies. O.Ka. and A.L. performed SPR studies. T.-M.M. produced antibodies for SPR. D.J.L., K.W.C., M.J.M., R.A.K., A.C., A.B.M., L.B.-F., A.S., J.R.P., R.E.W., A.S., J.B., A.M.R., W.H., D.R.A., S.M., F.R., A.L., V.P., D.S.L., A.T., D.M.B., M.R., J.M., O.K., N.K., J.B., D.D., planned, supervised, or carried out clinical trial activities or B cell sorting and sequencing assays or data organization leading to the data analyzed here. M.M.C and G.B.K.H. provided VH1-2 genotype analyses and quantification of mRNA and HCDR3 frequencies. J.R.W. provided VH1-2 allele inferences for post-vaccination BCRs. W.J.F., O.H., and A.C.D. performed statistical modeling and analyses. A.C.D., M.M.C, G.B.K.H., and W.R.S. wrote the main text and supplementary materials. A.C.D. and W.R.S. created figures and tables.

## Competing interests

W.R.S. and S.M are inventors on patents filed relating to the eOD-GT8 60mer immunogen in this manuscript.

## Data and materials availability

All data are available in the main text or supplementary materials.

## Notes

### Clinical Trial

NCT03547245

### Author Declarations

Institutional review boards at Fred Hutchinson Cancer Center (FHCC) and George Washington University (GWU) gave ethical approval for this work.

