## supplementary materials for "Human immunoglobulin gene allelic variation impacts germline-targeting vaccine priming"

Allelic variation in immunoglobulin heavy chain variable gene influences the efficiency of germline-targeting vaccine priming

Allan C. deCamp^1,†,^*, Martin M. Corcoran^2,†^, William J. Fulp^1,†^, Jordan R. Willis^3,4,5^, Christopher A. Cottrell^3,4,5^, Daniel M.L.V. Bader^3,4,5^, Oleksandr Kalyuzhniy^3,4,5^, David J. Leggat^6^, Kristen W. Cohen^1^, Ollivier Hyrien^1^, Sergey Menis^3,4,5^, Greg Finak^1^, Lamar Ballweber-Fleming^1^, Abhinaya Srikanth^6^, Jason R. Plyler^6^, Farhad Rahaman^7^, Angela Lombardo^7^, Vincent Philiponis^7^, Rachael E. Whaley^1^, Aaron Seese^1^, Joshua Brand^6^, Alexis M. Ruppel^6^, Wesley Hoyland^6^, Celia R. Mahoney^1^, Alberto Cagigi^6^, Alison Taylor^6^, David M. Brown^8^, David R. Ambrozak^6^, Troy Sincomb^3,4,5^, Tina-Marie Mullen^3,4,5^, Janine Maenza^1,9^, Orpheus Kolokythas^10^, Nadia Khati^11^, Jeffrey Bethony^12^, Mario Roederer^6^, David Diemert^12,13^, Richard A. Koup^6^, Dagna S. Laufer^7^, Juliana M. McElrath^1^, Adrian B. McDermott^6^, Gunilla B. Karlsson Hedestam^2,^*, William R. Schief^3,4,5,14,^*

Correspondence to: (A.C.D.), (G.B.K.H), and (W.R.S.)

**This PDF file includes:**

Fig. S1 to S3

Tables S1 to S13

Captions for Data S1 to S2

**Other Supplementary Materials for this manuscript include the following:**

Data S1 (Data_S1.xlsx)
Data S2 (Data_S2.xlsx)

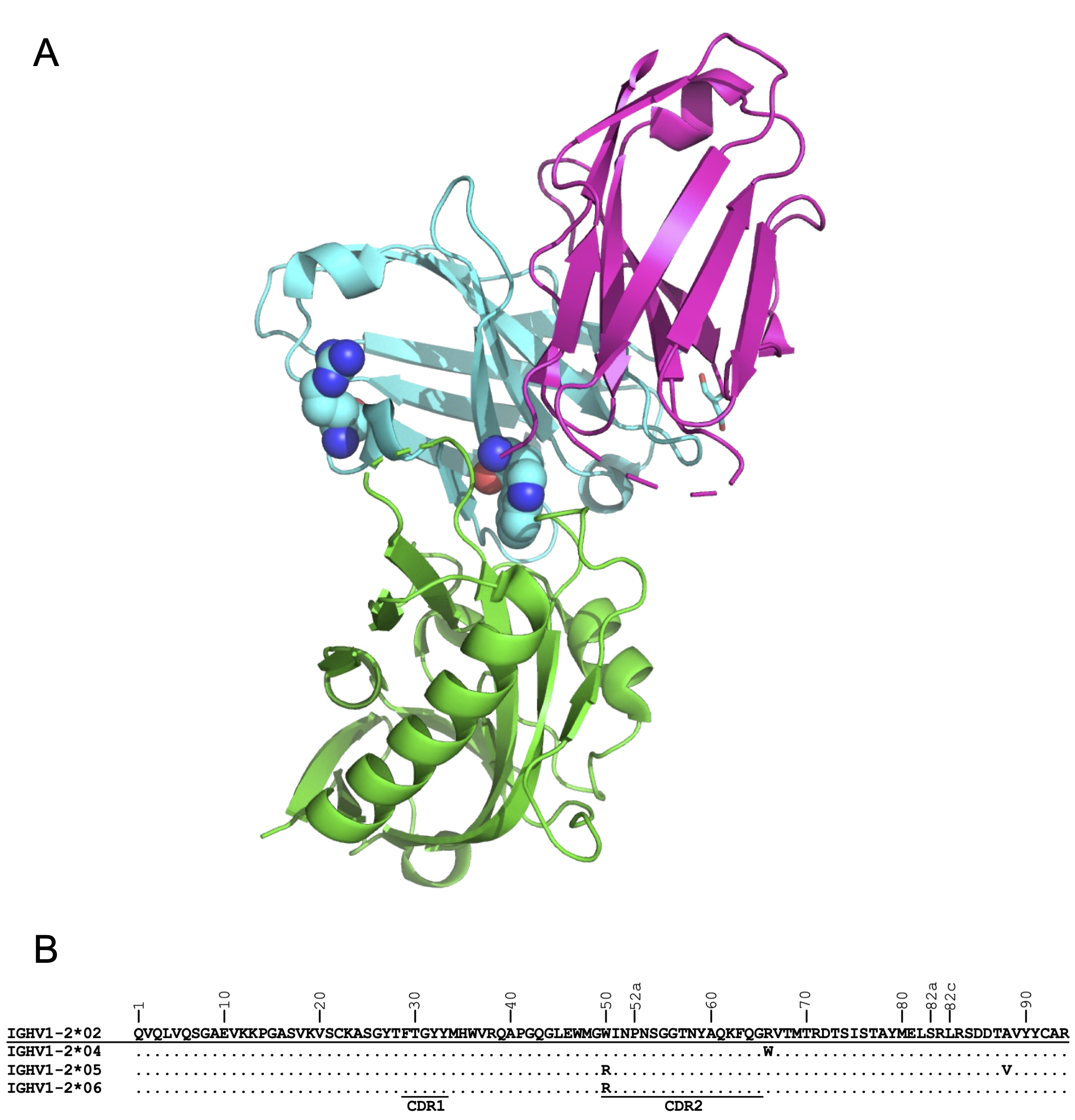

**Fig. S1. Locations of key allele-defining mutations in the structure and sequence of IGHV1-2. (A)** Structural model of eOD-GT8 (green) bound to a VRC01-class human naive precursor (cyan, purple), based on PDB: 5IES. Antibody heavy chain residues Trp50, present in IGHV1-2*02 but mutated to Arg50 in IGHV1-2*05 and *06, is shown in spheres at middle. Antibody heavy chain residue Arg66, present in IGHV1-2*02 but mutated to Trp66 in IGHV1-2*04, is shown in spheres at left. **(B)** IGHV1-2 sequence alignment showing alleles confirmed by single nucleotide polymorphism (SNP) analysis and other methods (*13, 44, 45*).

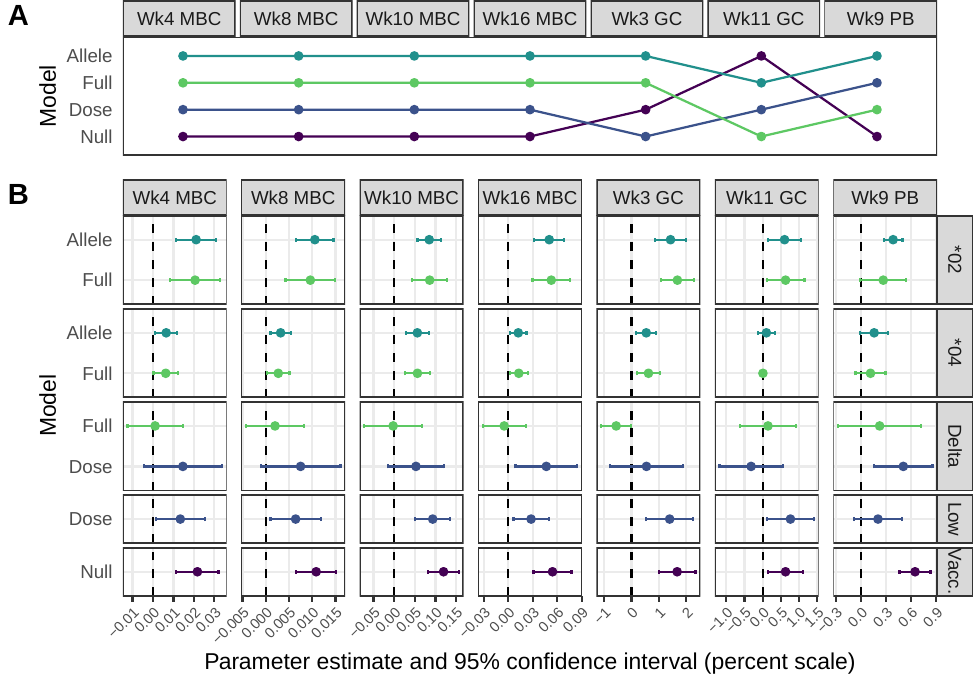

Fig. S2. Statistical modeling of post-vaccination VRC01-class B cell frequencies. (A) Model rankings from QAICc (see text and Methods) are shown as best to worst, from top to bottom, for each sample type and timepoint shown left to right. For all MBC timepoints, models were ranked in the same order from best to worst: (i) Allele, (ii) Full, (iii) Dose, and (iv) Null. Each color-coded line connects the ranking of the models across time points. (B) Points and lines show estimates and 95% confidence intervals (CIs) for each model parameter, for different models labelled on the y axis, at each time point labeled at the top of the graph. Model parameters representing the per-allele contributions of *02 and *04, the contributions of dose delta (100μg - 20μg) and dose=20μg, and a single estimate for the vaccine dose groups pooled (Null model) are grouped in rows *02, *04, Delta, Low and Vacc., respectively, as indicated on the right side of the panel. Parameter estimates and CIs are color-coded by model as in panel A and are displayed with values on the percent scale as indicated by the x-axis.

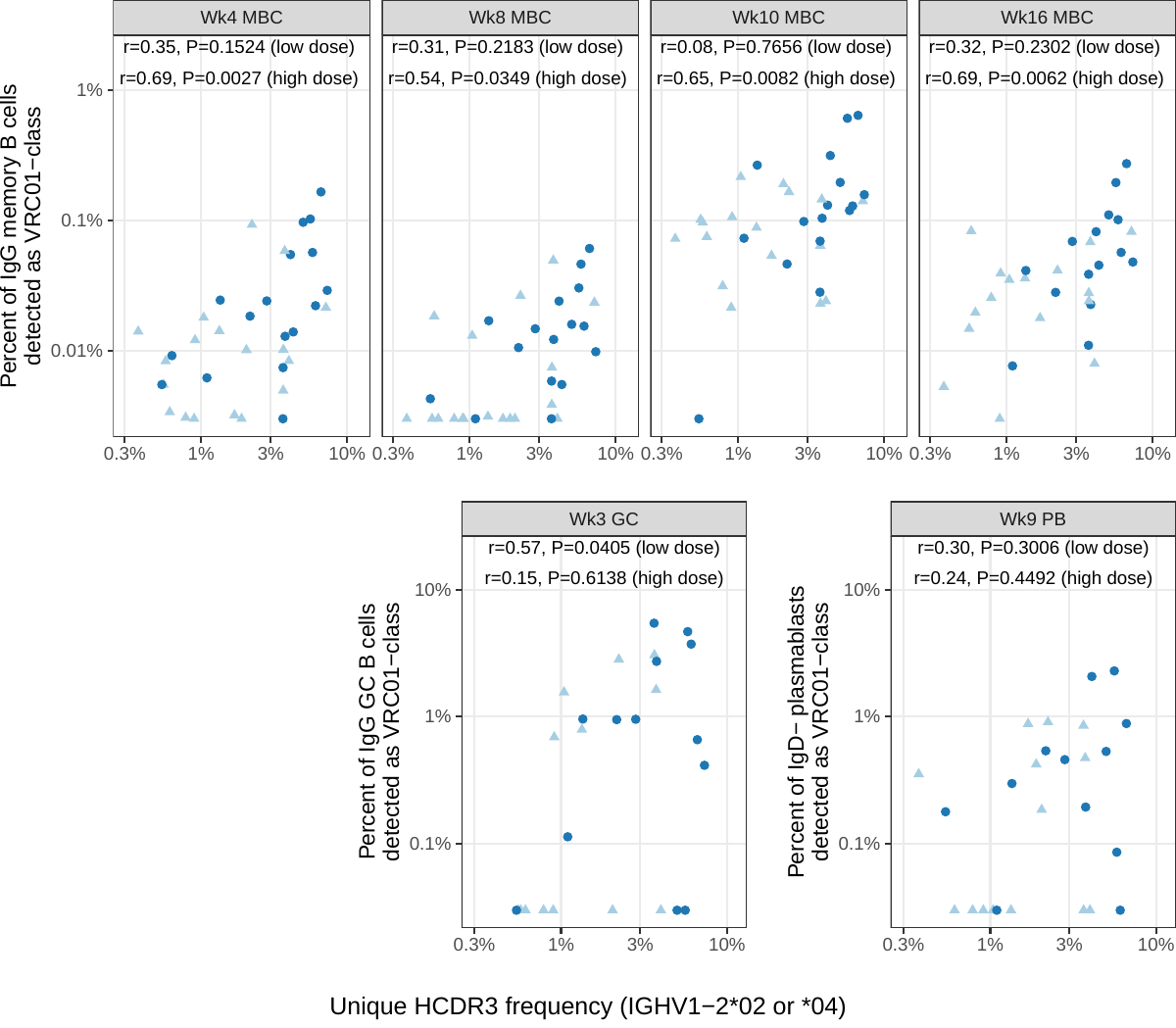

**Fig. S3. Correlations between pre-vaccination IgM unique HCDR3 frequency (IGHV1-2*02 or *04) and the percent VRC01-class B cell response by visit and treatment group.** Points are shape- and color-coded as shown in the legend. Spearman correlation coefficient (r) and P-values are displayed for each time point and dose group.

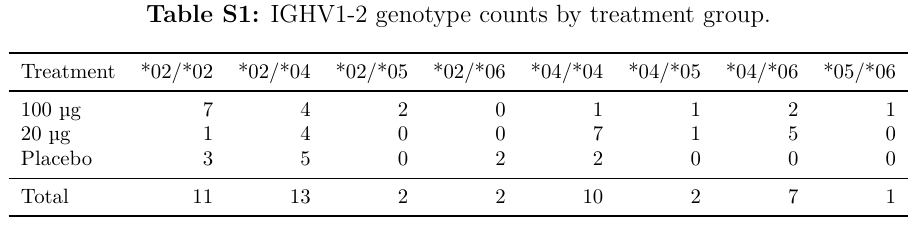

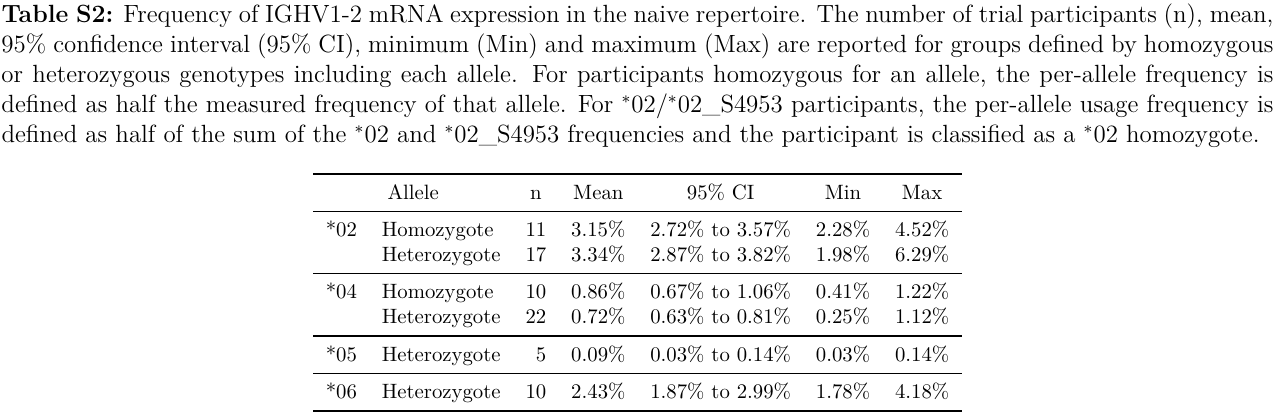

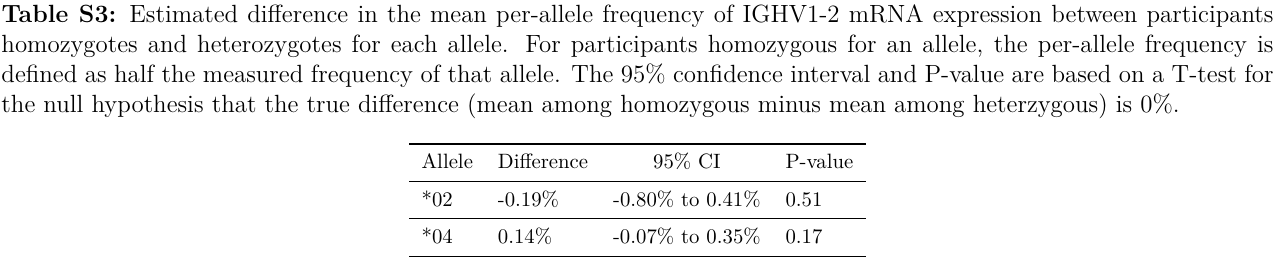

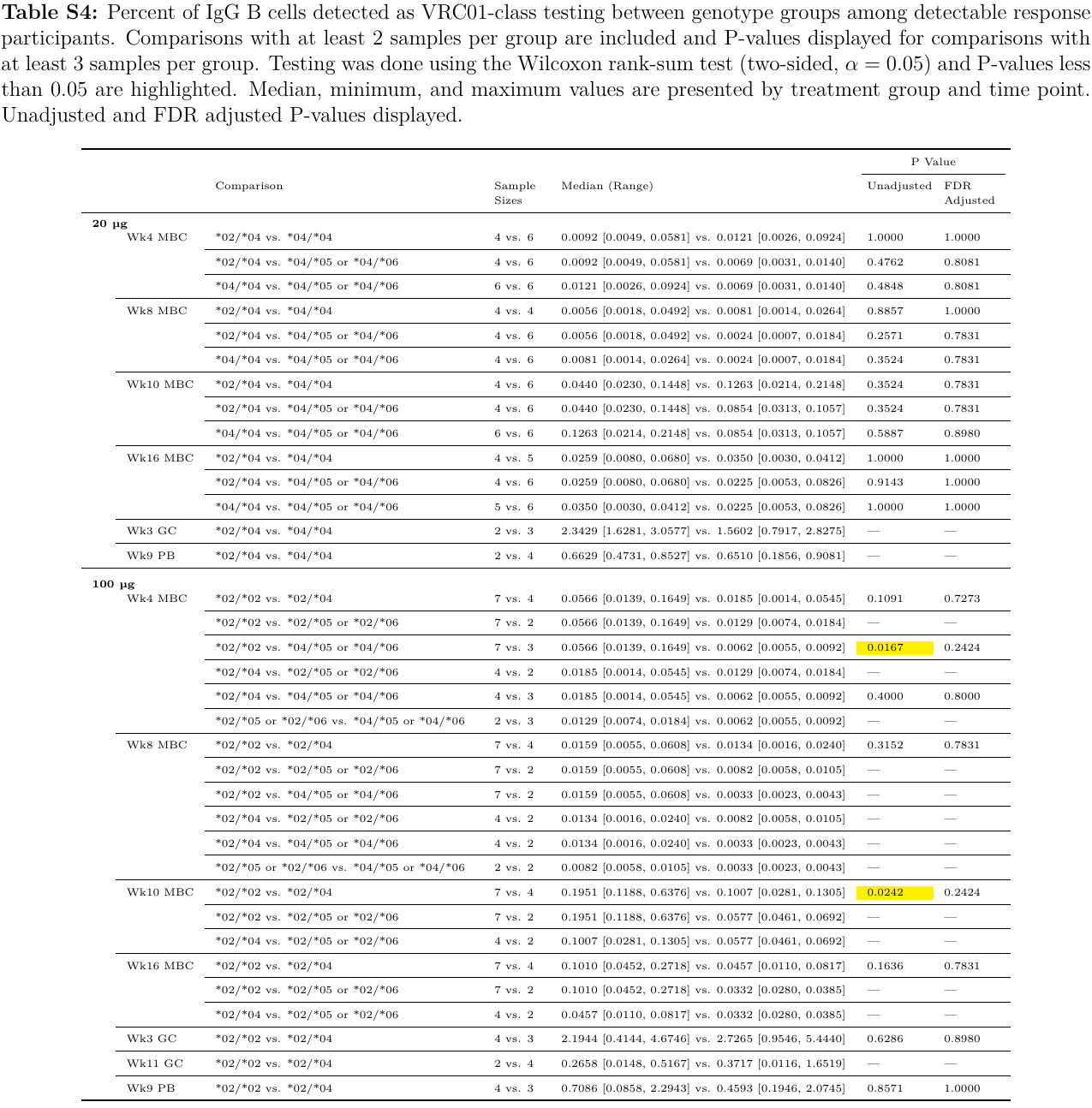

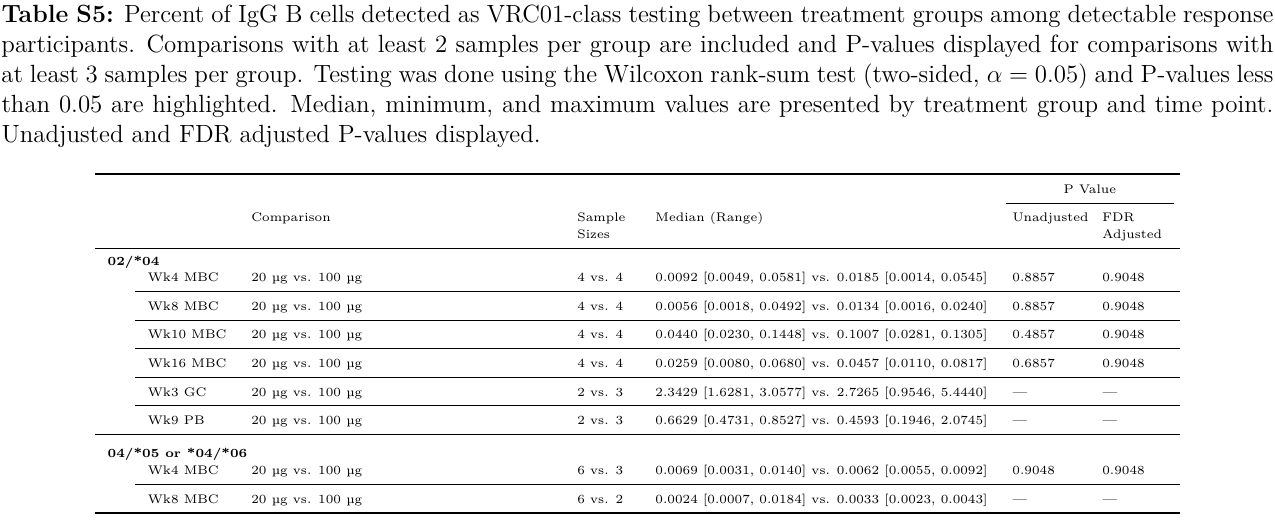

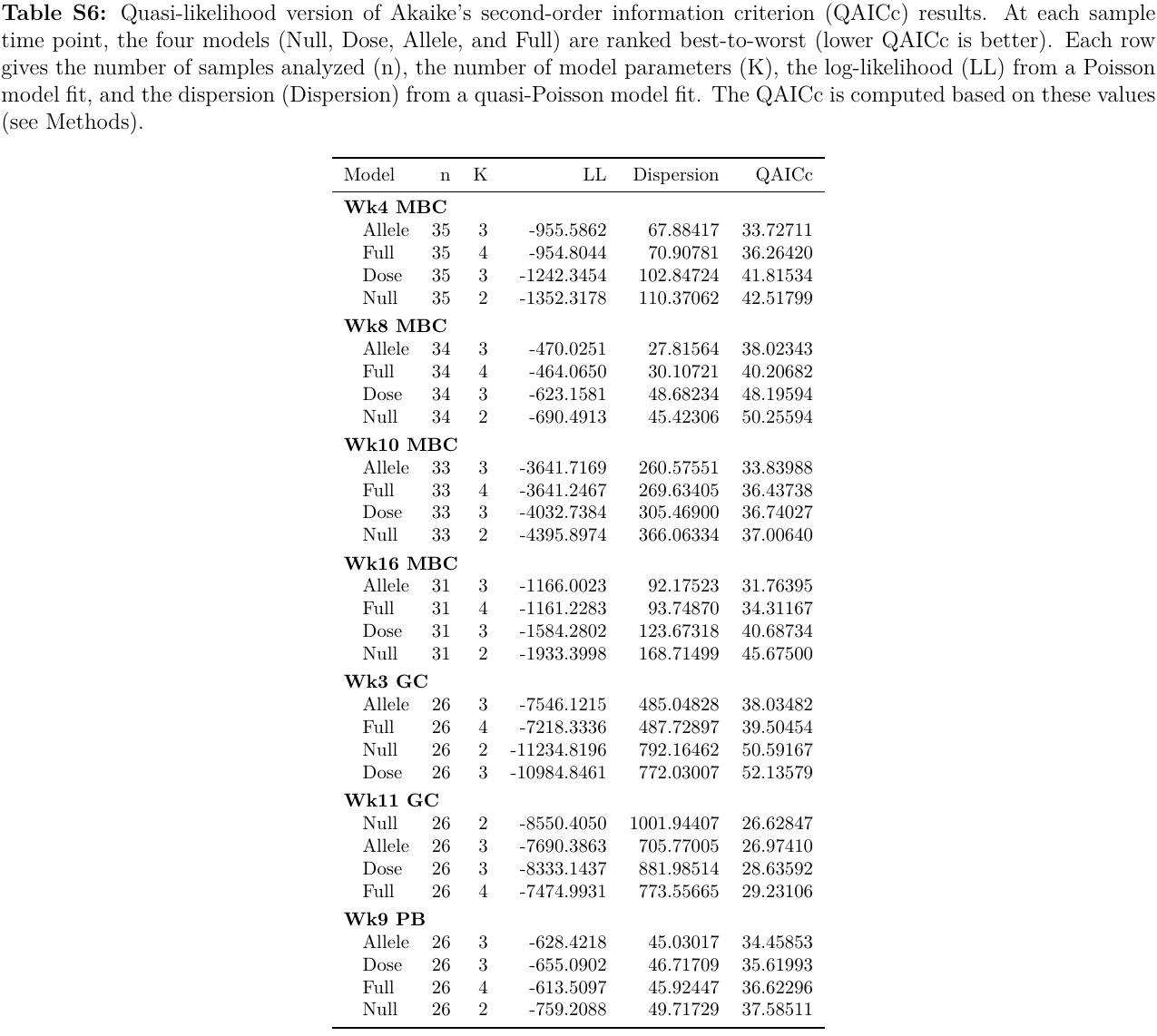

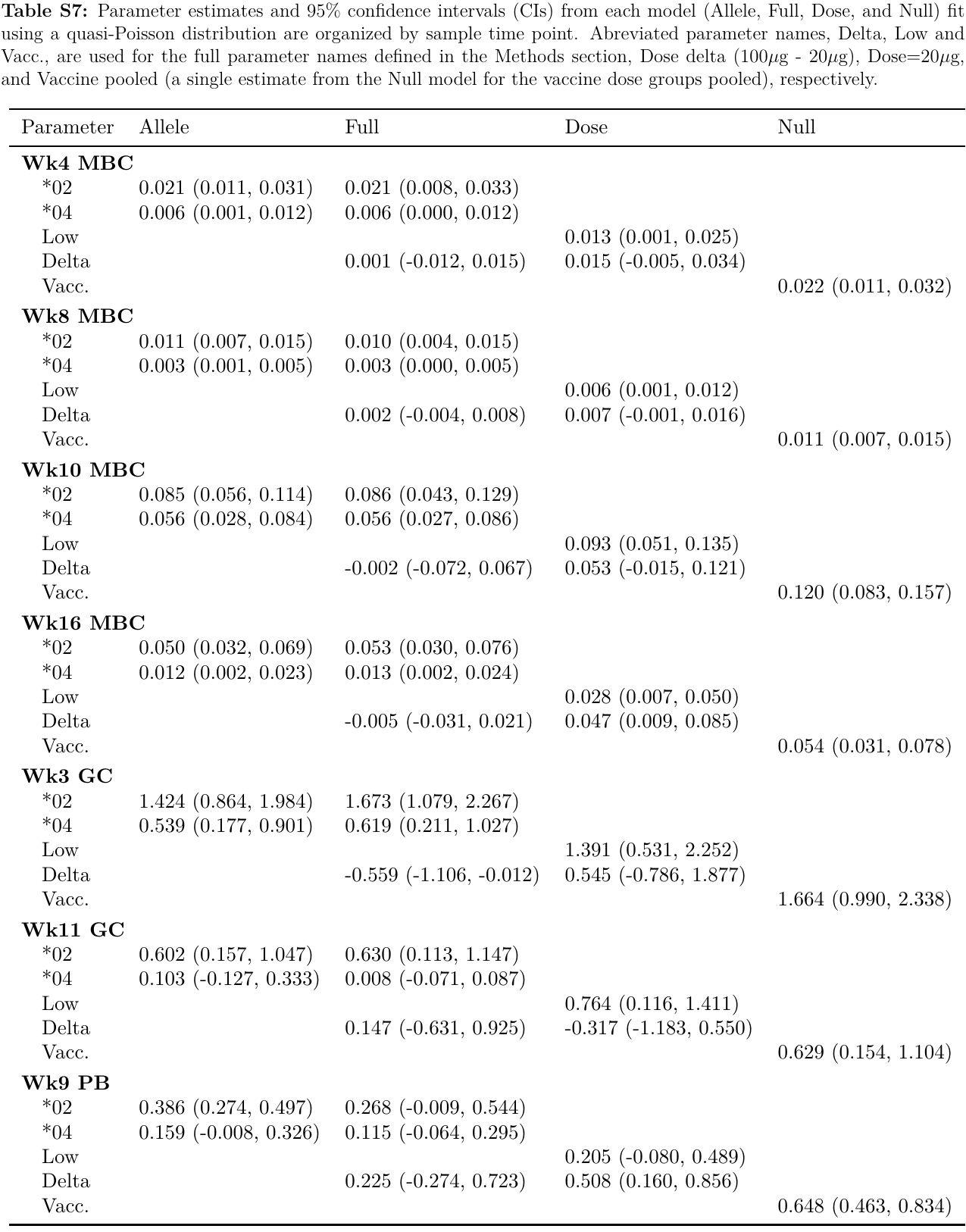

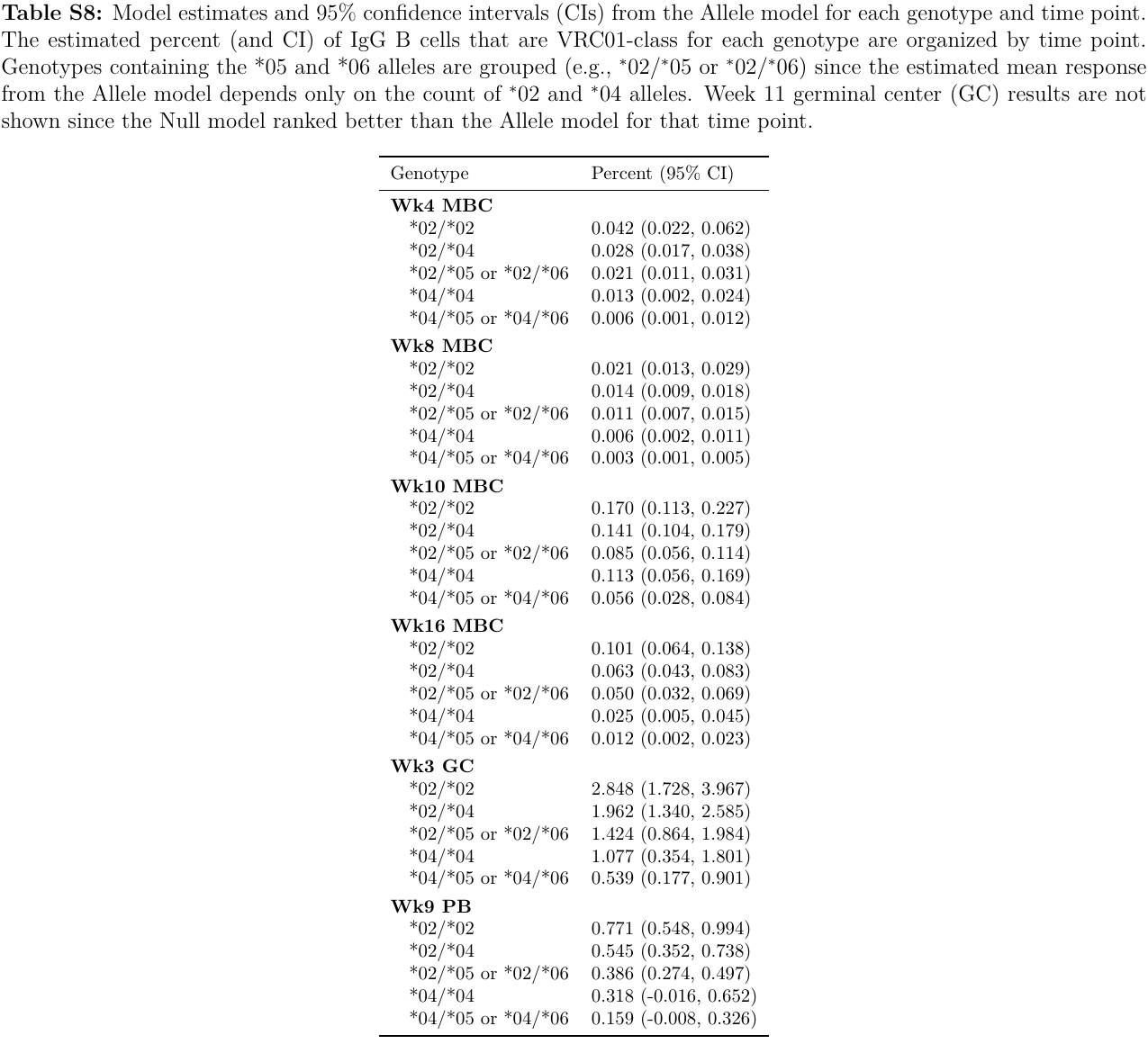

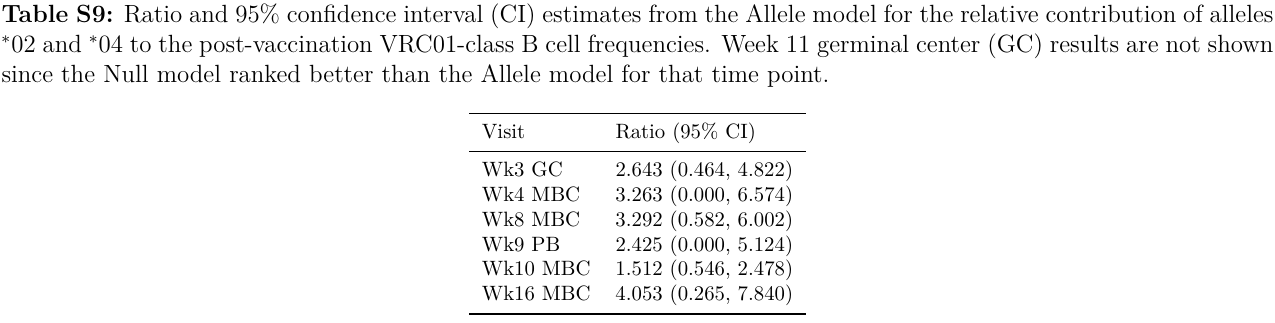

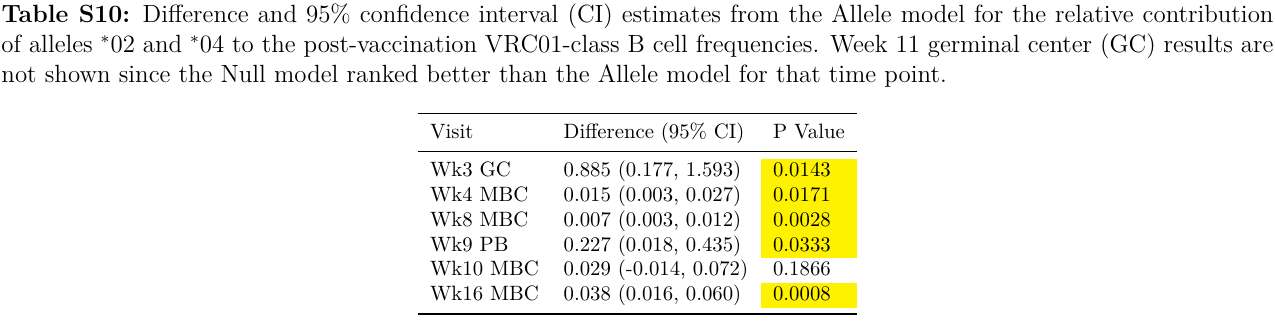

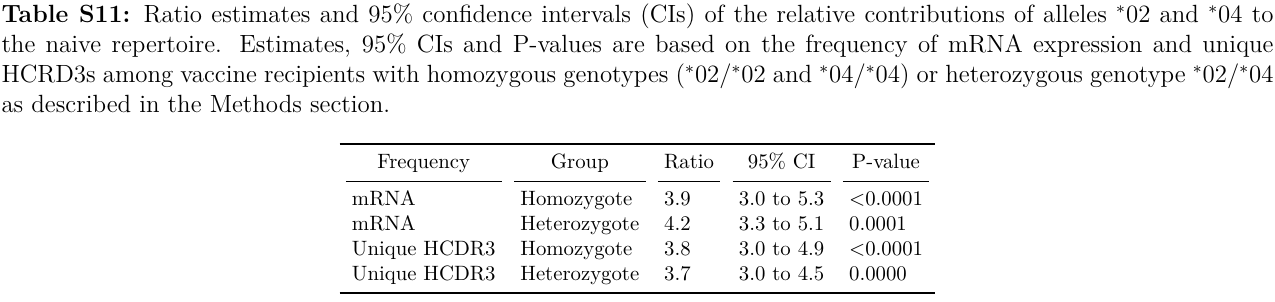

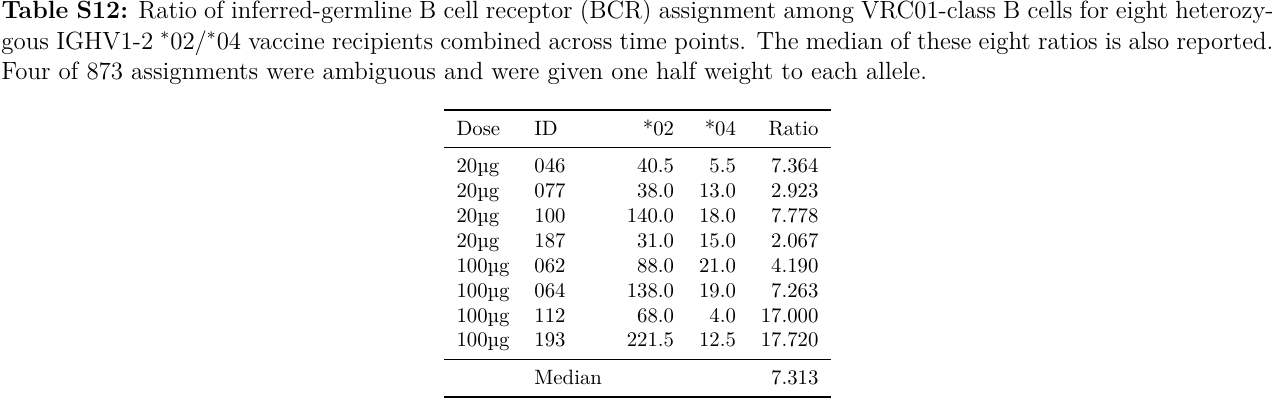

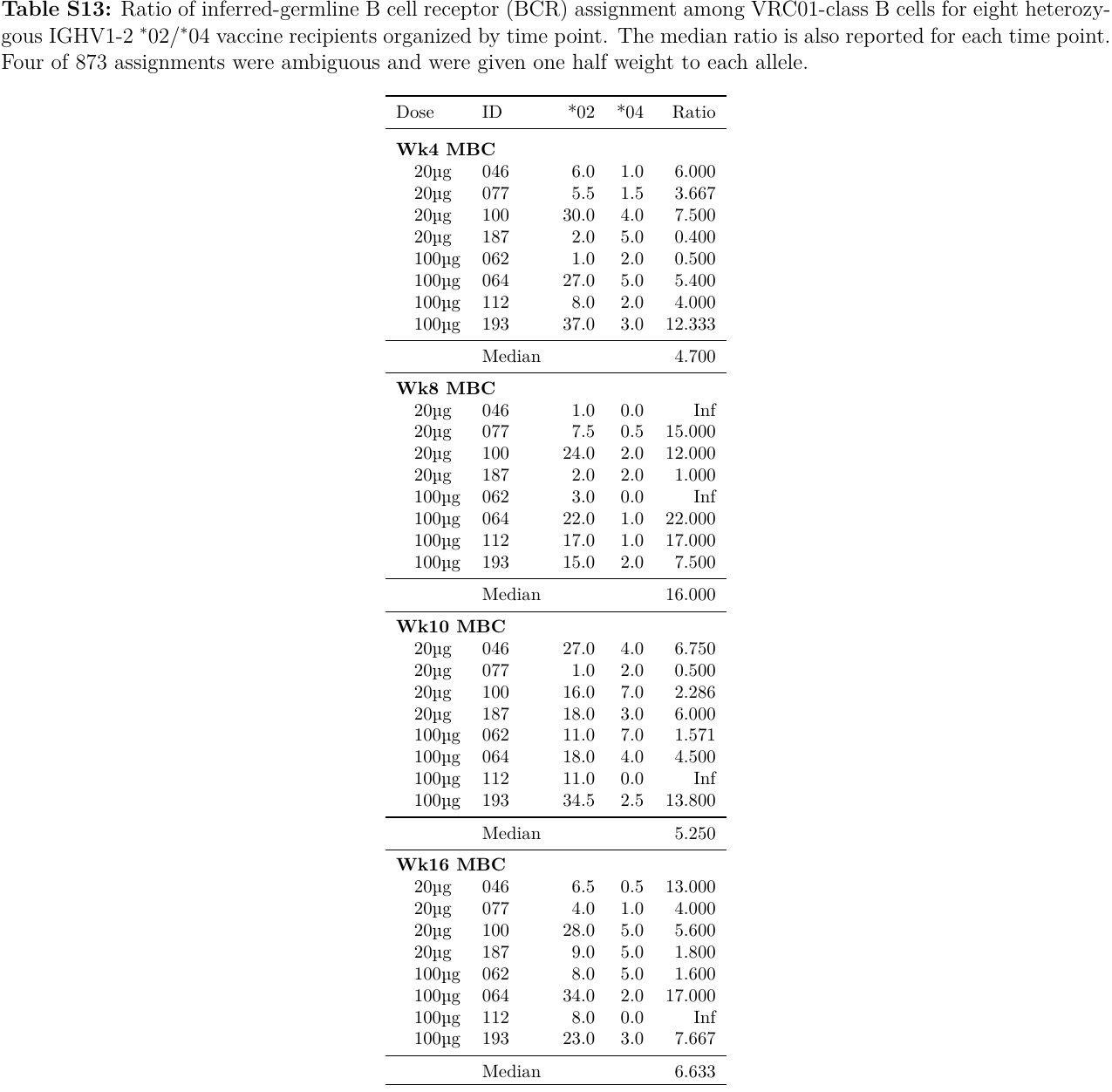

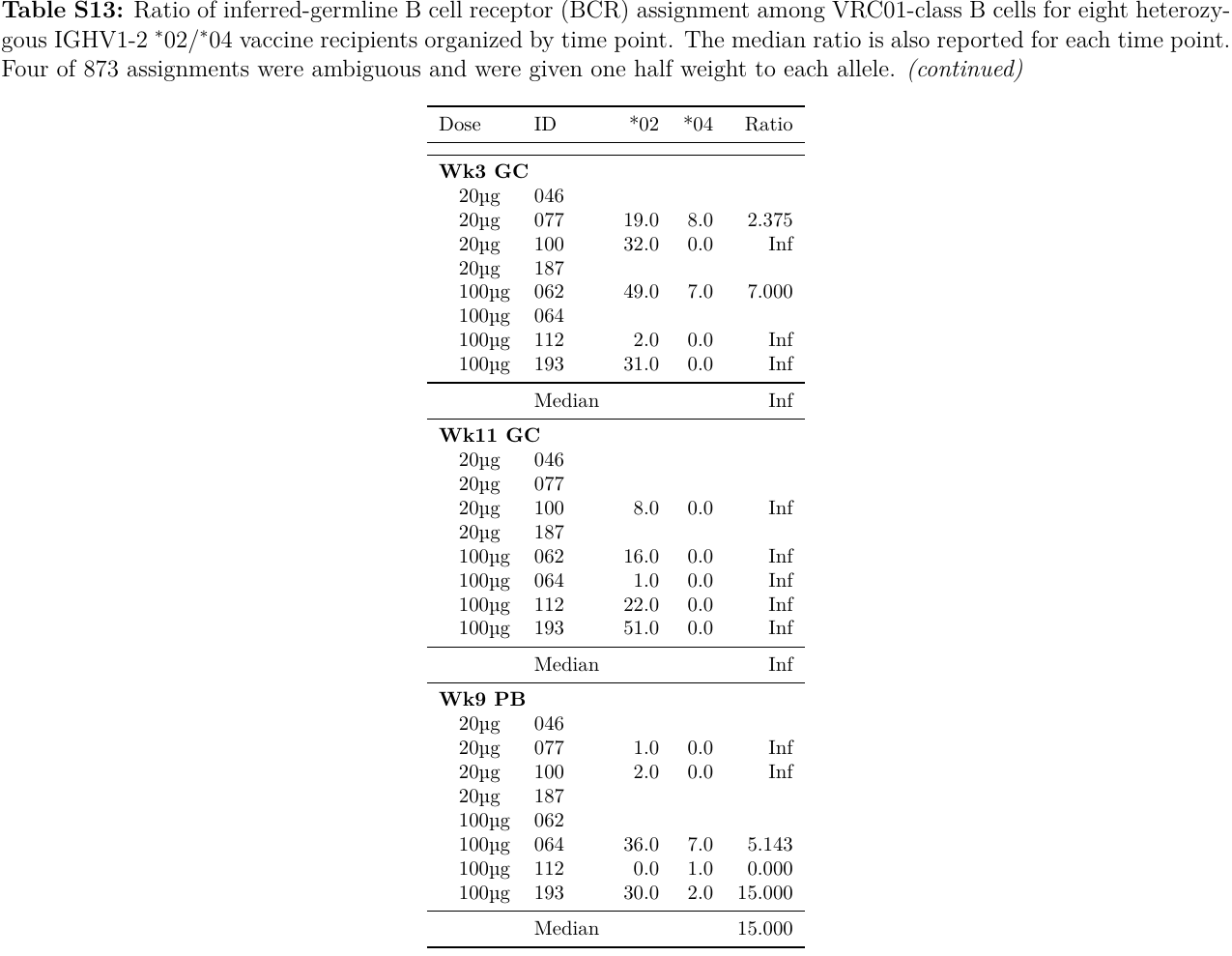

Data S1. (separate file)

Counts and frequencies for IGHV1-2 allele mRNA UMIs and unique HCDR3s in two IgM libraries for each trial participant.

Data S2. (separate file)

SPR data corresponding to Figure 5.
